# Implications of shared motor and perceptual activations on the sensorimotor cortex for neuroprosthetic decoding

**DOI:** 10.1101/2025.04.24.25326368

**Authors:** Alexander B. Silva, Jessie R. Liu, Vanessa R. Anderson, Cady M. Kurtz-Miott, Irina P. Hallinan, Kaylo T. Littlejohn, Samantha Brosler, Adelyn Tu-Chan, Karunesh Ganguly, David A. Moses, Edward F. Chang

## Abstract

Neuroprostheses can restore communicative ability to people with paralysis by decoding intended speech movements from the sensorimotor cortex (SMC). However, overlapping neural populations in the SMC are also engaged in visual and auditory processing. The nature of these shared motor and perceptual activations and their potential to interfere with decoding are particularly relevant questions for speech neuroprostheses, as reading and listening are essential daily functions. In two participants with vocal-tract paralysis and anarthria (ClinicalTrials.gov; NCT03698149), we developed an online electrocorticography (ECoG) based speech-decoding system that maintained accuracy and specificity to intended speech, even during common daily tasks like reading and listening. Offline, we studied the spectrotemporal characteristics and spatial distribution of reading, listening, and attempted-speech responses across our participants’ ECoG arrays. Across participants, the speech-decoding system had zero false-positive activations during 63.2 minutes of attempted speech and perceptual tasks, maintaining accuracy and specificity to volitional speech attempts. Offline, though we observed shared neural populations that responded to attempted speech, listening, and reading, we found they leveraged different neural representations with differentiable spectrotemporal responses.

Shared populations localized to the middle precentral gyrus and may have a distinct role in speech-motor planning. Potential neuroprosthesis users strongly desire reliable systems that will retain specificity to volitional speech attempts during daily use. These results demonstrate a decoding framework for speech neuroprostheses that maintains this specificity and further our understanding of shared perceptual and motor activity on the SMC.

## Introduction

Stroke or amyotrophic lateral sclerosis (ALS) can injure descending motor pathways, leading to paralysis and loss of the ability to move and articulate speech [1,2]. Recent work has demonstrated that neuroprostheses can restore naturalistic communicative ability to people with paralysis by recording brain activity and applying algorithms to decode this activity into intended movements and, thereby, speech [3–12]. These systems have predominantly leveraged representations of low-level movements from the sensorimotor cortex (SMC) that have persisted after paralysis [13]. However, the SMC also has neuronal populations activated by planning complex, coordinated movements [14,15] as well as processing visual and auditory stimuli [16–18]. While prior studies have identified “shared” neural populations, responding to at least two of these functions (e.g. speaking and listening [16,17,19]), it is an open question which areas of the SMC are most likely to contain these shared activations.

In the context of speech functions on the SMC, most work has studied only one or two of the three closely related communication modalities: speaking, listening, and reading. In addition to where shared populations are located in the SMC, the role of shared activity remains unclear. Some have hypothesized that SMC activations during speech perception are pre- or re-activations of representations of the corresponding vocal-tract movements [20–22]. However, others have proposed that shared activity is an important characteristic of neuronal populations important for motor planning of coordinated movements, like speech [15,16,23,24].

For speech neuroprostheses, the presence of shared activations raises two important concerns: could reading or listening unintentionally engage a speech neuroprosthesis? Relatedly, could simultaneous listening while speaking interfere with accurate decoding of speech attempts? Indeed, potential speech neuroprosthesis users strongly desire [25,26] systems that are only activated by volitional speech attempts and maintain reliable decoding performance in real-world settings where the user may switch between attempting to speak, in combination with reading, listening, or accessing mental imagery. In fact, a recent study found that their speech-decoding system was falsely activated when participants were engaged in a listening task [27]. Thus, there is a need to better understand the nature of shared activations on the SMC and to develop speech neuroprostheses that are robust to these non-target activations and specific to intended speech output.

Here, we leveraged a unique opportunity to investigate these questions in two participants with vocal-tract paralysis and electrocorticography (ECoG) arrays chronically implanted over their SMC. First, we asked whether a speech neuroprosthesis could be designed to ensure complete specificity to attempted speech even while reading or listening. We subsequently probed the localization and role of shared activity in our participants. We developed a speech-decoding system that maintained reliable decoding performance and had zero false-positive activations during listening, reading, or non-speech mental imagery. This was accomplished by training speech detection and verification models on cortical activity from reading and listening, in addition to attempted speech. Offline, we found that “shared” electrodes–those co-activated by attempted speech, reading, and listening–strongly localized to the middle precentral gyrus (midPrCG). Interestingly, these shared electrodes had distinct response profiles for each task, allowing them to distinguish between attempted speech and perception. Activity at shared electrodes was not strongly selective for different words during perception and these electrodes did not encode attempted vocal-tract movements during speech attempts. Instead, shared electrodes were selective for different words prior to speech attempts, suggesting a role in speech-motor planning, a necessary component for coordinated movement.

## Methods

Several methods follow procedures we have described in previous work (e.g. [9,28]), and are cited where appropriate. This study was conducted as part of the BCI Restoration of Arm and Voice (BRAVO) clinical trial (ClinicalTrials.gov; NCT03698149) approved by the FDA, UCSF IRB, and the NIH, with all ethical regulations rigorously followed in accordance with the Declaration of Helsinki. Although this data was collected as a part of this clinical trial, the data presented in this manuscript is not to inform concrete conclusions regarding the primary outcomes of the trial.

### Experimental design

The first participant (Bravo-1, he/him) was 30-40 years old at enrollment and had been diagnosed with severe spastic quadriparesis and anarthria by neurologists and a speech-language pathologist due to stroke of the bilateral pons [8]. His injury did not affect cognitive function and he has slight residual function of the vocal tract allowing for audible grunts and moans, though he is unable to produce intelligible speech. To communicate, he relies on an Augmentative and Alternative Communication (AAC) interface that utilizes residual head movements to spell out words.

The second participant (Bravo-3, she/her) was 40-50 years old at time of enrollment into the study and had been diagnosed with quadriplegia and anarthria by neurologists and a speech-language pathologist. Similarly, this was due to a large stroke of the pons with cognitive function left intact. She can vocalize a small set of monosyllabic sounds, such as ‘ah’ or ‘ooh’, but she is unable to articulate intelligible words. She similarly relies on an AAC interface that leverages residual head movements to facilitate spelling out words. After details concerning the neural implant, experimental protocols, and medical risks were explained, both participants provided full informed consent to partake in the study.

### Neural implants

Both participants were implanted with high-density electrocorticography (ECoG) arrays (PMT) and percutaneous pedestal connectors (Blackrock Microsystems). For Bravo-1, the array consisted of 128 electrodes with 4 mm center-to-center spacing implanted on the pial surface, covering a portion of the left frontal, precentral, and postcentral cortex (Fig. 1A). For Bravo-3, the array consisted of 253 electrodes with 3 mm center-to-center spacing implanted on the pial surface covering a portion of the left precentral, postcentral, and temporal cortex (Fig. 1A).

**Figure 1.**
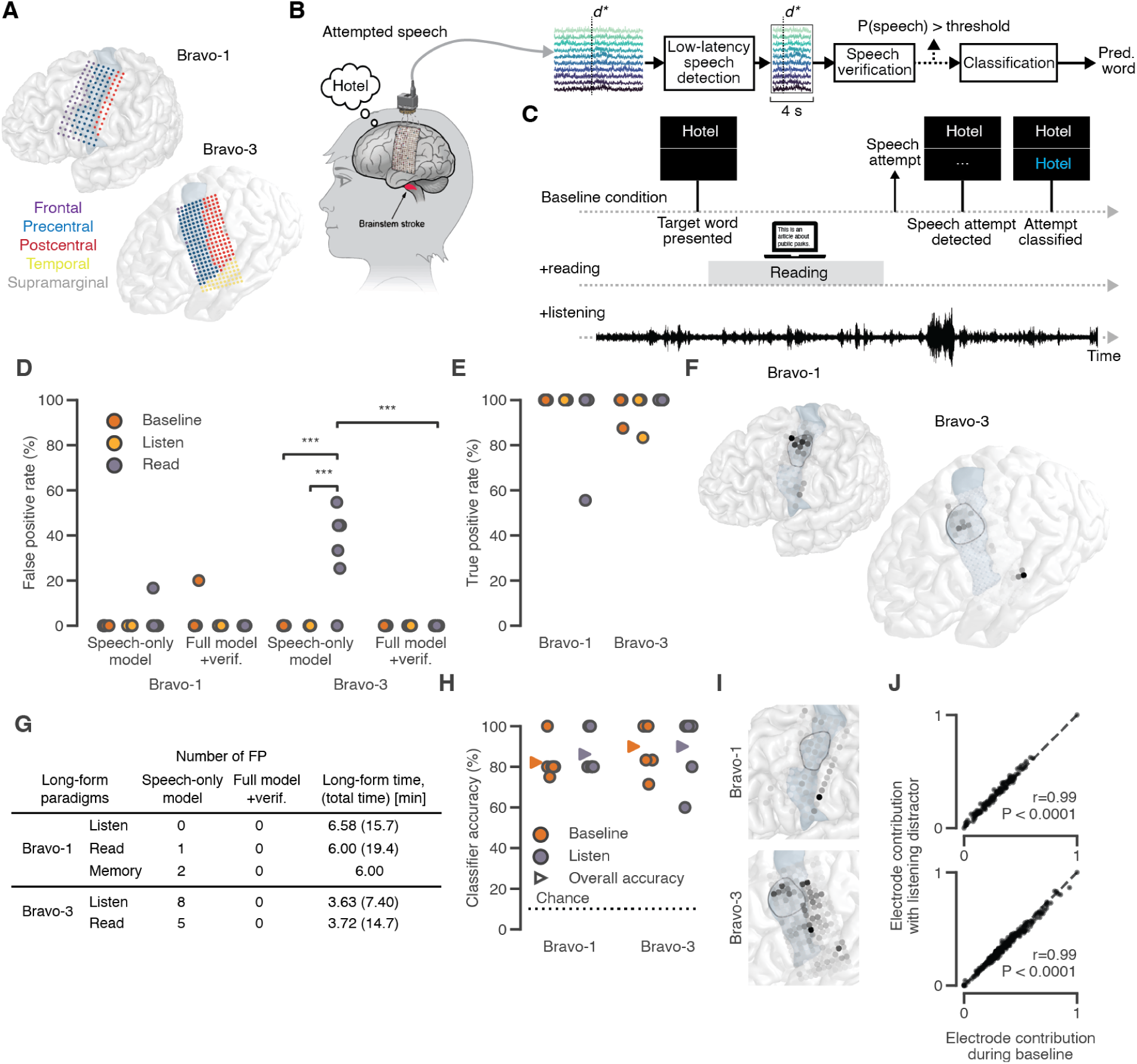
Reading and listening do not interfere with speech decoding. (**A**) ECoG grids in the two participants. Electrode color indicates anatomical region and the precentral gyrus is shaded. (**B**) The full speech-decoding framework used in the study (herein referred to as FS). d* denotes the detected onset of a speech attempt. The window around d* was [-0.5, 3.5] and [-1, 3] s for Bravo-1 and Bravo-3, respectively (**C**) The decoding framework was tested in three settings: “baseline”, “reading”, and “listening”. Across all, the participants attempt to speak at their leisure. (**D**) The false positive rate (FPR) of the FS versus a speech-detection model only trained on speech, across 6 pseudo-blocks for each condition. *** P < 0.0001 (Fisher exact test). (**E**) The true positive rate (TPR) of the FS across conditions. (**F**) Electrode contributions for the speech-detection model. (**G**) The number of false positives using the FS versus a speech-detection model only trained on speech during long periods of listening and reading (and “memory” during an autobiographical interview with Bravo-1) with no speech attempts. The amount of time where the system was active and could produce false positives is noted. The total amount of time with any one of the modalities (across long-form blocks and the main task including speech attempts) is in parentheses. (**H**) Accuracy of the 10-word classifier during baseline and listening trials across 6 pseudo-blocks. Offset triangle markers note the average accuracy across all trials. (**I**) Electrode contributions for the 10-word classifier. (**J**) Scatter plot comparing electrode contributions for the 10-word classifier during online evaluations with and without concurrent listening (Pearson correlation coefficient and p-value are noted). **(E,I)** MidPrCG outlined in black with the PrCG shaded blue.

Given that Bravo-3 was implanted after Bravo-1, newer hardware was available. Relative to the placement of the ECoG array in Bravo-1, placement of Bravo-3 was more posterior in order to be centered on the central sulcus and better capture relevant speech-motor representations [8]. The percutaneous connector transmits electrical measurements from the ECoG arrays to an external headstage (Blackrock Microsystems), where signals are digitized and sent to a computer for downstream analysis. Bravo-1 was implanted in February 2019 and Bravo-3 was implanted in September 2022. Online results for this project were collected in winter 2023-2024, 1752 days post implant for Bravo-1 and 492 days post implant for Bravo-3.

### Data acquisition and signal processing

We applied the same data acquisition and signal processing framework as in our previous work [4,28]. In brief, electrical field potentials from the ECoG contacts were common-average referenced with subsequent extraction of the high-gamma amplitude (HGA; 70-150 Hz) and low-frequency signal (LFS; 1-100 Hz) at each electrode at 200 Hz. A 30-second sliding-window z-score was used to separately normalize each ECoG channel’s HGA and LFS. Together, HGA and LFS are referred to as neural features and were used for both training and evaluation of online decoding models. All data collection and online decoding tasks were performed either at or near the participant’s residence. Decoding models were trained offline using NVIDIA Tesla V100 GPUs hosted on the lab’s server infrastructure. A custom Python real-time package [29] was used to coordinate data collection, task management, and online decoding.

### Artifact rejection

In Bravo-1, three trials were excluded due to an artifact affecting the quality of the HGA (Fig. S1). This artifact caused extended periods of decreased variance in the signal. We examined each trial, both qualitatively and quantitatively, to exclude trials affected by this artifact. Quantitatively, we computed the overall temporal variance across channels by first taking the standard deviation of the HGA for each electrode over the window [-5, 10] relative to every time point. Next, as an aggregate measure we took the 30th percentile of the HGA standard deviation across electrodes at each timepoint. A cut-off of 0.6 for this metric was used to identify trials affected by this artifact. Fig. S1 shows one of the three trials.

### Task design

To train and evaluate the speech-decoding system, we designed several different tasks.

### Isolated-target task

During the isolated-target task for attempted speech, the text for a target word appeared on the screen along with 4 dots on either side. Each dot sequentially disappeared followed by the text target turning green to indicate the participant should attempt to speak (go-cue). Bravo-1 attempted to speak with some residual vocalization while Bravo-3 silently attempted to speak (attempted miming). During the isolated-target task, we used a vocabulary of 10 total words (referred to as isolated words) for each participant. For Bravo-1 these words were a subset of the words used for a prior bilingual-decoding study [9] (Table S1). In Bravo-3 these words were a subset of the NATO-code words that represent the phonetic alphabet (Table S1) [4]. We used the same utterance sets to probe listening and reading. In these cases, one of the isolated words would either be played through the speakers or shown on the screen for the participant to read. For reading and listening, no countdown was used, and a version of the task where 10 sentences, rather than the isolated words, were shown was also collected. In the case of reading, we also collected data where false fonts (e.g. “δƱΦϞΨƟ”) were shown to the participant [30]. During the listening task, we kept volume as constant as possible across recording blocks. To ensure that listening was not strongly modulated by volume, in the small range of volumes used for data collection, we quantified the correlation between maximum HGA and volume level for each electrode. Overall, we found no positive significant correlations (Fig. S2; Spearman correlation). As an additional control, we had both participants attempt non-speech movements. The task was identical to the attempted-speech task except the 10 words were replaced with various non-speech movements: flex each of the 10 fingers, raise and lower the eyebrows, blink eyes, swing head side to side, shrug each shoulder, swing each arm out, curl each arm, bend each wrist, swing each leg, curl each-side toes, and bend forward.

Neural features collected during the isolated-target task were used to train and optimize deep-learning models trained to detect and verify attempted speech as well as classify content of attempted speech, reading, and listening.

### Online-decoding task

The goal of the online-decoding task was to test the robustness of the speech-decoding system to perceptual tasks. In this task, one of the isolated words appeared on the screen. The participant could then attempt to speak this word whenever desired. After a speech attempt was detected, verified, and classified (Fig. 1B), the screen would be cleared of all text for 4 s before the next trial would begin with a new isolated word on the screen. The task was collected in blocks where each of the 10 isolated words was presented once. We collected this task under three conditions (Fig. 1C). In the first “baseline” condition, no perceptual task was present. In the second “listening” condition, a podcast was consistently playing throughout the task and the participant took time to listen to the podcast before making a speech attempt. Finally, in the third “reading” condition, a laptop opened to an online article was placed in front of the participant (the experimenter would scroll based on a head cue from the participant). The participant took time to read portions of the article before making each speech attempt. For Bravo-1, he spent an average of 24.8 ± 13.7 s per trial (mean ± standard deviation) reading portions of the article (with a range of 9.29 to 59.8 s across all trials). For Bravo-3, she spent an average of 19.7 ± 7.05 s per trial reading portions of the article (with a range of 8.12 to 35.2 s across all trials).

Speech detection, verification, and classification models were trained offline prior to online-system use with the isolated-target task. Speech detection models were trained to identify attempted speech events from the neural features. Speech verification models were trained to classify whether an isolated-target trial was reading, listening, or attempted speech. Classification models were trained to output the identity of the word based on attempted-speech events.

### Modeling

The overall speech-decoding system consisted of three deep-learning models. The first was designed to run continuously and detect speech at low latencies. The second was trained to verify a detected speech event as being attempted speech, and not reading or listening. The third was trained to classify detected speech events as one of the isolated words. As depicted in Fig. 1B, the speech-detection model first identifies a detected speech onset (d*). The speech-detection model then waits for a 3 or 3.5s window of neural features after detected onset, to define a detected event. The time after detected onset is necessary for the classification model, as in prior work [8]. The detected event is then passed to the verification model and, if the verification model accepts the event, the event is subsequently passed to the classification model. Overall, latency is most strongly incurred by waiting for a 3 or 3.5s window after detected onset whereas inference latency of the verification model is on the order of a hundred ms using cpu. All model hyperparameters and weights were chosen/trained based on task data from data-collection sessions prior to online evaluations and no recalibration occurred during online evaluations. While no day-of recalibration occurred during online testing, for Bravo-3 online evaluations with the reading condition occurred over a month after evaluations with the listening condition. In this case, all models were retrained on data collected during the intermediate month.

### Speech detection

The speech detection model was designed and trained similarly to our prior works [8,9,28]. In brief, the speech detection model consisted of 3 unidirectional long short-term memory (LSTM) layers followed by 1 fully connected layer. The model continuously processed incoming neural features at 200 Hz and generated continuous probabilities over 5 classes–silence, preparation, speech, reading, and listening. These probabilities were then processed to generate discrete windows corresponding to detected speech attempts. The processing consisted of smoothing the probabilities with a causal moving average window, thresholding the smoothed probabilities to generate a binary signal, and then a time threshold to determine the onset and offset of attempted speech events. Relevant model parameters are detailed in Table S2.

For training, labels corresponding to each of the 5 target classes were manually created following the same general principles, with some adjustments per participant. For the isolated words task, time points between the presentation of the word on the screen and the go-cue were labeled as “speech preparation”. Time points from the go-cue to some offset from the go-cue (for Bravo-1, to 2.75 s after go-cue, for Bravo-3, to 85% of the trial duration) were labeled as “speech”. Time points from this go-cue offset to the end of the trial (before the screen cleared for an inter-trial interval) were not used during training as it is ambiguous when each participant had finished the speech attempt and returned to rest. These offsets were chosen based on qualitative observations of each participant’s behavior. All other time points were labeled as “silence”. For the listening task, all time points during the stimulus presentation were labeled as “listen” and the remaining time points as “silence”. For the reading task, all time points when the target word was on the screen were labeled as “read” and the remaining time points as “silence”. In addition to the listening and reading versions of the isolated-words tasks, each participant also listened and read a set of 10 phrases. These were labeled in the same way. Additionally, 1 minute-long blocks were collected with each participant where they were instructed to rest. All time points in these blocks were labeled as “silence” and used during training.

For Bravo-1, model performance qualitatively appeared to be better when not including reading in the training set, and so no reading data was included. Additionally, several test runs of the online decoding task (baseline condition) infrastructure were piloted with Bravo-1. As these blocks had no explicit go-cues, we manually annotated the acoustic onset and offset of his speech attempts, labeled them as speech and all other time points as silence, and used this during training. Online evaluation blocks were collected on a single day, with the exception of the long memory-reflection block which was collected 48 days later. The same online model was used in both cases.

For Bravo-3, various phrase-level data was being collected alongside the aforementioned datasets. For convenience, more recent phrase-level data was labeled according to the methods stated above and used as the primary source of attempted speech examples. Online evaluation blocks were collected on two separate days, 43 days apart. For the first day, the online speech-detection model was trained with the phrase-level speech-attempt data, 10-phrase and 10-word listening data, rest data, and 10-word reading data (10-phrase reading data was accidentally excluded). For the second day, the same dataset plus 10-word speech attempt data and 10-phrase reading data was also used to retrain a new model.

In all cases, models used for online evaluation were only trained on data prior to that day.

### Speech-detection ablations

In order to assess whether including examples of reading and listening helped reduce any potential false positives, we also trained versions of the speech-detection model that did not use reading or listening data and evaluated them offline on the evaluation blocks. For Bravo-1, a single model was retrained, which used a subset of the data used to train the online model (e.g. just removing listening data). For Bravo-3, two models were retrained as two models were used for online evaluation. The first model was trained using a subset of the data that the online model used on the first day of online evaluations–excluding reading and listening data, however also including prior isolated words data. The second model was similarly trained using a subset of the data used to train the online model used on the second day of evaluations. These models were used to offline evaluate the blocks from the first and second day, respectively. No model parameters were changed for these ablations.

### Latency of speech-detection model

In order to generate discrete detected events that correspond to speech attempts, continuously predicted probabilities (at 200 Hz) go through aforementioned post-processing. While smoothing and probability thresholding are causal or do not impose latency, time threshold does impose a latency as it must wait for the smoothed predicted speech probability to be above a certain threshold (the probability threshold parameter) for at least a certain number of time points (the time threshold parameter). Similarly, the probability must return to below this threshold for at least a certain number of time points to determine the offset of the event. In this paper, we set those parameters to 100 time points, or 0.5 s. If detected events were perfectly aligned to the onset and offset of speech attempts, there is still a 0.5 s latency before the onset and offsets are each recognized, yielding a consistent 0.5 s latency to detecting an onset or offset in addition to the latency of those from the onset or offset of the true speech attempt.

For Bravo-1, we could measure the onset and offset of his speech attempts by annotating his vocalizations. We found that there was a median 0.14 s onset latency and a median 0.89 s offset latency. For Bravo-3, since she silently attempted to speak, we could not similarly compute the latency based on vocalization time.

Though in this work, we detect discrete events and classify them, the speech detection model is amenable to lower latency frameworks either by reducing the time threshold parameter or using the continuously predicted speech probabilities at 200 Hz.

### Speech verification

Speech verification models were trained with the objective to classify a detected speech event as being either attempted speech, listening, or reading. To train such a model, we used attempted speech, listening, and reading data from the isolated-target task. For reading and listening, we included neural features from both isolated words and sentences (false fonts were not included) aligned to the onset. For attempted speech data, rather than aligning to the go-cue, we aligned to the detected onset of each trial (see above, ‘Speech detection’). For Bravo-1, we collected a few practice online-decoding blocks a week before testing and included these trials in training. In all cases, we trained the speech-verification model on a window of [-0.5, 3.5]s and [-1, 3]s around the trial onset for Bravo-1 and Bravo-3 respectively. Similar to prior work [4,28], the model itself consumed neural features at a rate of 33.3 Hz (downsampled by a factor of 6 from 200 Hz) and consisted of a one-dimensional temporal convolutional layer followed by 2 gated recurrent neural network (GRU) layers with a final fully-connected layer that produces probabilities over 3 classes: attempted speech, reading, and listening. During model training, we used stochastic gradient descent and the Adam optimizer [31] to find a set of parameters that minimized the cross entropy loss between target and predicted outputs. During online testing, any event with an attempted-speech probability of over 0.65 was verified to be associated with attempted-speech. This was chosen based on probability distribution from the isolated-target task. A table containing the precise hyperparameters used is provided in Table S3.

### Classification

Classification models were trained on the attempted-speech data from the isolated-targets task and a few practice online-decoding blocks (as in ‘Speech verification’) for Bravo-1. Neural features were aligned to detected onsets for each trial and a window of [-0.5, 3] s and [-1, 3] s was used for Bravo-1 and Bravo-3 respectively. The classification architecture and process was the same as previous work [4,28]. Neural features were first downsampled by a factor of 6 before being passed through a one-dimensional temporal convolutional layer and a series of GRU layers (2 for Bravo-3, 3 for Bravo-1). A fully-connected layer of output dimension 10 was used to map the final latent state from the GRU layers onto a distribution over the isolated words. The softmax function was used to scale this output to a probability distribution. Similar to above, model training was guided by stochastic gradient descent and the Adam optimizer to find a set of parameters that minimized cross entropy loss between target and predicted outputs. For both online classification and speech-verification, 10 randomly initialized models were fit on the training data to improve prediction accuracy via ensembling. A table containing the precise hyperparameters used is provided in Table S4.

### Evaluation

To evaluate the speech-decoding system in the presence of perceptual interference, we collected 9 total blocks of the online-decoding task: 3 baseline, 3 with reading, and 3 with listening. This gave a total of 30 trials for each condition with 3 repeats of each isolated word. In Bravo-3 a fourth baseline block was collected, giving 40 total trials. In Bravo-1, the three trials affected by the artifact were excluded (Fig. S1). Additionally, to further probe the specificity of our system to attempted speech over longer time scales, we collected online-decoding blocks where the participant listened to a podcast, read articles, or performed a mental imagery task for several minutes (see Fig. 1L). During these blocks, however, the participant was instructed to not make any speech attempts. We confirmed with the participants that they were actively reading and listening during the corresponding online-decoding blocks. Correspondingly, we found that many of the electrodes activated by listening in the isolated-target task also had significantly increased HGA during the longform listening blocks (100% in Bravo-1, 79% in Bravo-3). This same trend was seen for reading (97.4% in Bravo-1, 95.2% in Bravo-3). To test whether HGA was increased during the longform blocks we sampled 100 random time points in the longform and baseline online decoding blocks. We computed the average HGA in the surrounding 4 s window and then applied Wilcoxon rank-sum tests between average HGA in the baseline versus listening or reading longform blocks. Holm-Bonferroni correction was applied across electrodes within each participant.

### True positive and false positive calculations

The true positive rate (TPR) is defined as the proportion of true speech attempts that were correctly detected by the decoding system while the false positive rate (FPR) is defined as the proportion of detected events that do not correspond to a true speech attempt. To identify true and false positive detected events, we first manually annotated all true speech attempts. For Bravo-1, this was done based on the recorded audio while for Bravo-3, this was done from face camera recordings (as she silently attempts to speak while Bravo-1 audibly attempts to speak). If a detected event is within 0.5 s of a true speech attempt, it is assigned to that speech attempt and would be considered a true positive. If one or more detected events meet this criteria for a single speech attempt, they are all assigned to that speech attempt. This does not affect the calculation of TPR, as that speech attempt is still counted as a true positive only once. For the calculation of FPR, only 1 of the detections associated with that speech attempt is counted. We chose to do this because including both detections would increase the denominator and could make the FPR appear better than it may truly be.

### Offline characterizations of the speech-verification model

To further characterize the speech-verification model, we used the online evaluation blocks for Bravo-1 and Bravo-3. As a first characterization, we varied the speech-probability threshold between 0 and 1 and computed the corresponding false positive and false negative rate (as a proportion of total evaluation trials) using the online speech-verification models. As a second characterization, we restricted the context length of time windows around detected onsets that were passed to the speech-verification model. We then computed the accuracy of the model in accepting or rejecting the online detected events, using the speech-probability thresholds found optimal offline (Fig. 1G). This allowed us to probe speech-verification performance as a function of context length following a detected event.

### Pseudo-blocking trials from online results

Since there were 30 (and for one condition 40 for Bravo-3) trials for the online results for each condition, we used a pseudo-blocking method in order to visualize distributions of TPR, FPR, and 10-word classification accuracy. We generated 6 pseudo-blocks per condition (therefore an average of 5 trials per pseudo-block), and then calculated the TPR, FPR, and classification accuracy for each pseudo-block. These 6 points were used for visualization Fig. 1 and Fig. S3. Though pseudo-blocks were created for visualization purposes, they were not used for statistical analysis (see Statistics section).

### Offline cross validation of classification models

Similar to prior work [9], to evaluate the performance of speech-verification and classification models across different regions, we used a cross validation approach over all the isolated-target data. In brief, we repeated the above-described training procedure, using neural features only from electrodes in a given region. We evaluated the models using 10-fold cross validation (CV). In each of the 10-folds, 90% of the data is used for training and 10% for evaluation. Among the 90% of data used for training, an additional 10% of that is reserved as a validation set, used to define when to stop neural network training. Within each of the 10 folds, 5 randomly initialized models were fit to ensemble predictions over the held out evaluation fold.

### Neural signal analyses

To measure activation in response to attempted-speech, reading, and listening at each electrode, we used the data from the isolated-target task. Instead of a running 30s z-score, used for online decoding, we z-scored each trial to the silent, inter-trial period before stimulus presentation. For each task, we then measured responsiveness at every electrode based on Wilcoxon rank-sum tests comparing the average HGA during the pre-stimulus window to the average HGA after onset of the function ([0,1]s for attempted speech and reading and [0.5,1]s for listening). For each function, p-values across the 253 electrodes were corrected for multiple comparisons with the Holm-bonferroni method and a cutoff of 0.05 was used for statistical significance. For visualization of task-modulated activation, the Z-statistic at each electrode was visualized with negative statistics (indicating higher baseline activity) set to 0. To compare electrodes with shared activations to those that encoded articulatory movements, we used our prior published estimates of supralaryngeal encoding for Bravo-3 (methodology for how these values were computed can be seen in Metzger et al. 2023 [4] ).

For low-frequency band analyses, power spectral density was estimated on the isolated-target data during attempted speech, reading, and listening using a multi-taper method [32] in the theta (5-12 Hz) and beta (20-30 Hz) ranges. For every trial the mean power in these ranges was computed for the 0-2 s window relative to task go-cue or onset. Trials were then pseudo-blocked by taking the mean over consecutive, non-overlapping groups of 60 trials. Distributions across each task were then z-scored for each band.

### Electrode contributions

For speech detection, speech verification, and 10-word classification models, electrodes contributions were calculated by first taking the derivative of the classifier’s loss function with respect to the input features over time [4,8,33]. For each electrode, the L2-norm was computed over the time-dimension of the derivative. This resulted in a single magnitude per electrode per time course. For speech verification and 10-word classification models, these time courses were for each trial. For speech detection, these time courses were over each block. These values were then averaged across time courses yielding a single contribution score per electrode. For speech detection, electrode contributions corresponding to the HGA features are plotted in Figure 1. Given that LFS substantially improved classification performance in prior work [28], for classification and speech verification, this process was performed separately for HGA and LFS features and for each electrode the contribution score is defined by the sum of the two. The electrodes contributions for the classification models in Figure 1 are plotted on a log scale to better visualize the overall distribution of contributions.

### Statistical analysis

All statistical evaluations were performed using nonparametric tests and noted alongside significance statements in figure captions and the results section. For the online blocks, because there were much fewer trials, we used the Fisher exact test to compare between conditions for each participant for the TPR, FPR, and classification accuracies. For offline results on the isolated target data, where there are many more trials, Wilcoxon Rank-sum tests are used. Associated p-values for Spearman rank or Pearson correlation coefficients were computed via permutation testing (1000 iterations). For any multiple comparisons, p-values are corrected (within participants) using Holm-Bonferroni correction. Boxplots in all panels depict median (horizontal line inside box), 25th and 75th percentiles (box), 25th and 75th percentiles ±1.5 times the interquartile range (whiskers) and outliers (diamonds).

## Results

We developed a speech-decoding system that maintained accuracy and specificity to intended speech during common daily tasks like reading and listening. We studied two right-handed participants (ClinicalTrials.gov; NCT03698149) with vocal-tract paralysis and anarthria, referred to as Bravo-1 and Bravo-3. In both, cortical activity was measured with electrocorticography (ECoG) arrays covering frontal, precentral, postcentral, and temporal cortex in the left hemisphere (Fig. 1A). Bravo-1 and Bravo-3 were implanted in February 2019 and September 2022, respectively. Bravo-3’s ECoG array is higher density (253 versus 128 electrode contacts) and extends into the temporal lobe. Bravo-1 attempted to speak, producing audible grunts that were unintelligible. Bravo-3 silently attempted to speak, making uncoordinated vocal-tract movements. To decode cortical activity into intended speech with maximal specificity, even during perceptual functions, we designed a system consisting of three key components (Fig. 1B). While each participant attempted to say a target word from a predefined 10-word vocabulary, we streamed the high-gamma activity (HGA; 70-150 Hz) and low-frequency signals (LFS; 0.3-100 Hz; see Methods) from each electrode to a speech-detection model trained to identify volitional speech attempts at low latencies (approximately 0.5 s, see Methods). Next, windows around speech events from the detector, -1 to 3 s for Bravo-3 and -0.5 to 3.5 s for Bravo-1, were passed to a speech-verification model, trained to disambiguate attempted speech from reading and listening. This model acted as an additional mechanism to maximize specificity and, if the probability of attempted speech exceeded an acceptable threshold, the speech event was passed to a classifier, which outputs the most likely word.

### High specificity of the speech neuroprosthesis across conditions

We next simulated common daily-use scenarios of the neuroprosthesis, evaluating specificity to intended speech when the participants switched between attempting to speak and reading or listening. In each block, a target word was first presented on the monitor and the participant attempted to speak the word at their own pace (Fig. 1C). This attempt was detected, verified, and classified by the system, after which the task moved to the next trial. In the baseline condition, no perceptual content was present during the block. In the reading condition, the participant read sections of an article on a separate laptop before making a speech attempt (Video S1). Finally, in the listening condition, a podcast was playing for the duration of the block, and the participant listened to the podcast before making a speech attempt (Video S2).

An ideal system would have a zero false positive rate (FPR), defined as the proportion of detected events that are not associated with any speech attempt, and maximize the true positive rate (TPR), defined as the proportion of true speech attempts that are detected. For Bravo-1, FPR was not significantly increased across conditions (Fig. 1D, P > 0.01, Fisher exact test), when using the full system (depicted in Fig.1B and referred to as “Full model + verif.”). There was only one false positive detection, which was associated with a laugh, for an overall FPR of 1.15%. Notably, of Bravo-1’s 31 involuntary non-speech vocalizations (characteristic of his paralysis), only one resulted in the aforementioned false positive. For Bravo-3, FPR was unaffected and remained at 0.0% for all conditions. For both participants, we found that the presence of listening or reading did not significantly decrease the TPR (Fig. 1E, P > 0.01, Fisher exact test), which was 95.6% and 98.0% overall for Bravo-1 and Bravo-3, respectively.

For the full system, two primary design choices were made to maximize specificity: (1) including examples of listening/reading in training the speech detector (for Bravo-1, only listening was added, see Methods) and (2) including the speech-verification model. While the speech detector works at lower latencies, the speech-verification model receives a larger window relative to the onset of each detected event (4 s; Fig. 1B), leveraging longer context lengths to differentiate attempted speech from reading and listening. Offline, we assessed the effects of these design choices on TPR and FPR. Removing both the speech-verification model and using a speech detector trained only on speech attempts (and not reading or listening examples), significantly increased the FPR during the reading condition for Bravo-3 (P < 0.0001, Fisher exact test, Fig. 1K, Fig. S3A) but not during the listening condition. For both participants, neither design choice significantly affected the TPR, indicating the increased specificity did not come at the expense of rejecting true speech attempts (Fig. S3B). Neural activity in the midPrCG contributed most for detecting speech attempts in Bravo-1. For Bravo-3, both the midPrCG and a small cluster in the superior temporal gyrus (STG) were most important for detecting speech events (Fig. 1F).

To further challenge the system’s specificity, we recorded a handful of longer trials (roughly 2 minutes each) while the participants were asked not to make any speech attempts and only listen (Video S3), read, and, for Bravo-1, perform a mental imagery task where he was instructed to think of various memories via autobiographical questions. Using the full system online (“Full model + verif.”), we observed zero false positives for a total duration of about 18.6 and 7.35 minutes of reading and listening for Bravo-1 and Bravo-3, respectively (Fig. 1G). When combined with the conditions described above (Fig. 1C), this amounts to no false positives over 41.1 and 22.1 minutes of reading and listening for Bravo-1 and Bravo-3, respectively (Fig. 1G). Additionally, in Bravo-1 there were no false positives over 6 minutes of a mental imagery task, demonstrating robustness to a form of internal thoughts. Removing both the speech-verification model and using a speech detector trained only on speech attempts increased false positives, across longform conditions, to rates as high as a false positive every 27.2 s (Bravo-3) and every 180 s (Bravo-1). Thus, both training the speech detector on reading and listening data (Fig. S3C-D) and having an explicit speech-verification model (Fig. S3E; P < 0.01, one-sided Wilcoxon signed-rank test) can increase system specificity.

In this study, the classification model waits for 3 (Bravo-3) or 3.5 (Bravo-1) seconds of neural features after a detected onset (Fig. 1B). The speech-verification model is run over the same window, only adding minimal inference latency on the order of 100 ms. However, in future systems, the model making predictions (i.e. a classifier or a continuous decoder [4,6,34]), may not require 3-3.5 s of neural features after detected onset, which means a verification model could add significant latency. We simulated speech-verification performance when the context-length after the detected onset was reduced. In Bravo-1, 95% of detected events are correctly accepted or rejected with reduced false-positives as quickly as a second after detected onset. However, in Bravo-3, performance rose more slowly, reaching above 95% with no false positives around 2 s (Fig. S4). Latencies of 1-2 s do not strongly affect conversational discourse [35,36] and the improved specificity from the verification model (Fig. S3E) may be preferred by end-users. To better understand when users prefer added specificity at the expense of (1-2 s) latency, we surveyed our participants’ preference during daily-use scenarios. Across all settings, Bravo-1 preferred a high-specificity, but 1-2 s latency system, whereas Bravo-3 preferred this system in formal settings, such as a work presentation. For informal settings, she strongly preferred a system with negligible latency and some false positives (Table S5). Though only in two participants, these results highlight (1) the utility of both high and low latency methods to minimize false positives and (2) the advantage of a customizable speech-decoding system. Our system is flexible as the speech-verification model can be removed and the threshold at which a speech-event is accepted can be adjusted based on preference for false positives and false negatives (Fig. S4).

Through offline testing, we also confirmed that performance of the speech detection and verification models generalized to larger vocabulary sizes. We used a vocabulary of 104 English and Spanish words [9] and 1024 English words for Bravo-1 and Bravo-3, respectively. Importantly, since prior speech detection models were not assessed during reading and listening, we compared the large-vocabulary rates to rates computed from baseline blocks in our study. We found comparable large-vocabulary FPRs of 3.05% and 1.28%, relative to the FPRs in this study of 3.57% and 0.00%, for Bravo-1 and Bravo-3, respectively. We also found comparable large-vocabulary TPRs of 97.47% and 99.44%, relative to the TPRs in this study of 100.00% and 99.44%, for Bravo-1 and Bravo-3, respectively. Similarly, we retrained the speech-verification model on the same larger vocabulary datasets in Bravo-1 and Bravo-3. We found comparable large-vocabulary accuracy of 87.7% and 98.5%, relative to the accuracy in this study of 86.0% and 98.3%, for Bravo-1 and Bravo-3, respectively.

During the listening condition with attempted speech (Fig. 1C), the audible podcast may also affect the system’s classification accuracy. To test this, we compared classification accuracy between the baseline condition and the listening condition. We did not include the reading condition as reading was never concurrent with speech attempts whereas the podcast was audible for the entire block, including during speech attempts. Classification accuracy was 82.1% without and 86.2% with the listening condition for Bravo-1, and 90.0% for both for Bravo-3, amounting to no significant differences between the two conditions (P > 0.01, Fisher exact test, Fig. 1H). In contrast to the speech detector, vSMC electrodes along the central sulcus contributed most to the classifier (Fig. 1I). The classifier placed importance on the same electrodes during the baseline and listening condition (Fig. 1J; Pearson correlation, r = 0.99 and P < 0.0001 for both Bravo-1 and Bravo-3).

### Spatial and spectrotemporal characteristics of shared activity

We next characterized the nature of shared activations on the SMC and the features that enabled differentiating between reading, listening, and attempted speech. For each electrode, we tested whether HGA was significantly increased during each of the three tasks, relative to pre-trial resting activity (Fig. 2A-C; one-sided Wilcoxon rank-sum tests with 128-way (Bravo-1) or 253-way (Bravo-3) Holm-Bonferroni correction). Across our two participants, “shared” electrodes activated across reading, listening, and attempted speech, localized primarily to the midPrCG (Fig. 2A). Bravo-3 had a higher number of shared electrodes than Bravo-1 (42 versus 4), potentially explaining the higher false-positive rates seen without proper design considerations in Bravo-3 (Fig. 1D,G). Interestingly, shared electrodes did not overlap with electrodes that encoded attempted vocal-tract (speech articulatory) or hand movements (Fig. 2B). Aligning with posterior shifts in motor activation after stroke [37], electrodes encoding specific movements were primarily on the postcentral (PoCG) [38] and posterior precentral (PrCG) gyri, over the central sulcus. Across both participants, no electrodes were responsive during reading and not also during attempted speech (Fig. 2C). A majority of reading electrodes were also responsive during listening (Fig. 2D). Of note, we observed fewer electrodes responsive to listening in the midPrCG of Bravo-1 than Bravo-3, aligning with variability seen across individuals [39]. As expected, listening responses were also found in the temporal lobe [40–42] and overlapped strongly with attempted-speech but less so with reading (Fig. 2D-E).

**Figure 2.**
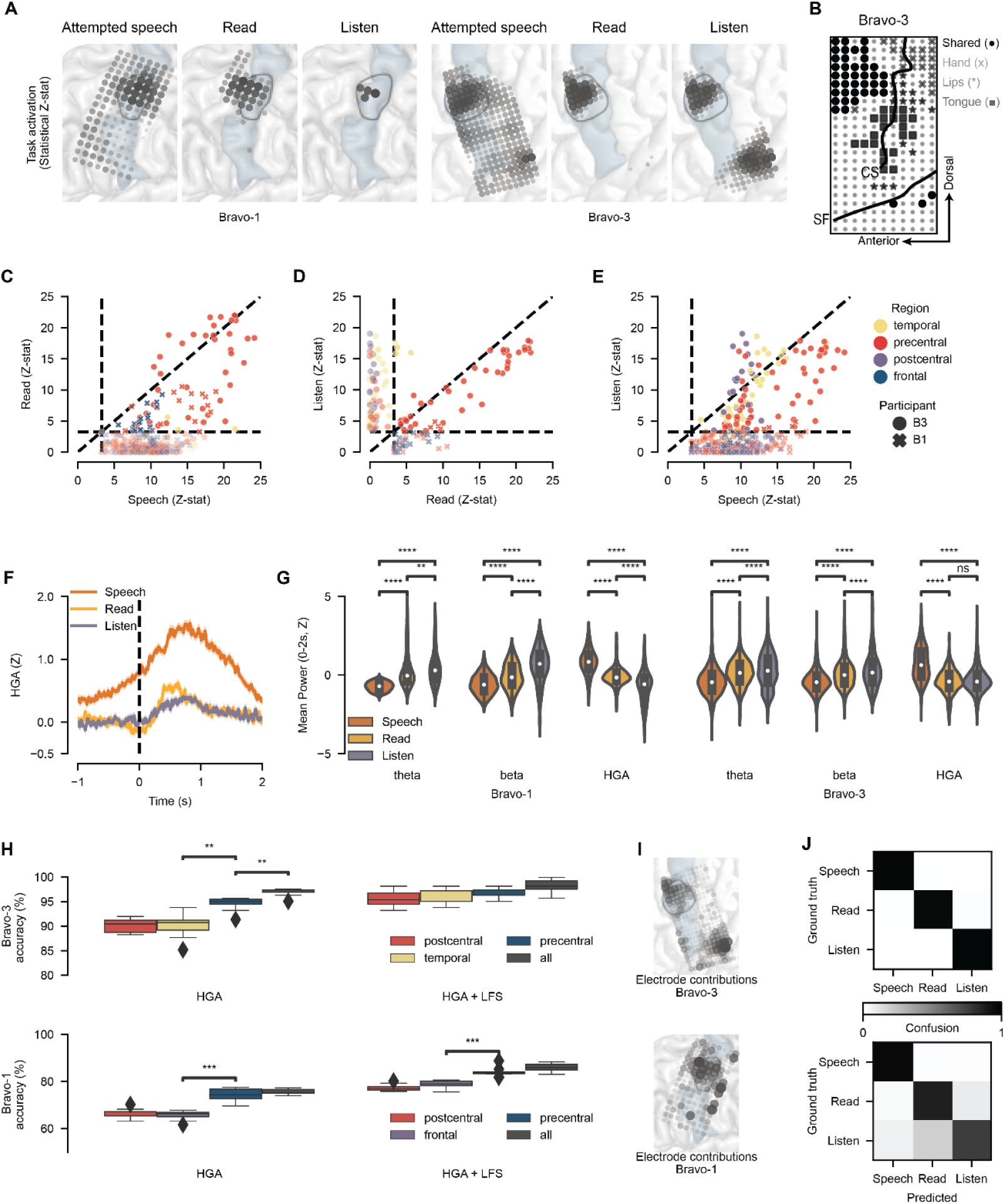
Shared and distinct cortical activations for reading, listening, and attempted speech. (**A**) Electrode responsiveness measured by non-parametric tests of pre-trial versus post go-cue mean high-gamma amplitude (HGA). Electrodes with significant task modulation (P < 0.05; one-sided Wilcoxon rank-sum tests with 128-way (Bravo-1) and 253-way (Bravo-3) Holm-Bonferroni correction) are shown. Results for Bravo-3 (**right)** and Bravo-1 (**left).** (**B**) In Bravo-3, shared electrodes, active for reading, listening, and attempted speech (calculated as in **A**), are visualized alongside electrodes encoding movements of the vocal-tract articulators and hand. Lines signify annotations of the central sulcus (CS) and sylvian fissure (SF). (**C-E**) One-sided Wilcoxon rank-sum test statistics (as in **A**) for each electrode plotted for reading and speech (**C**), speech and listening (**D**), and listening and reading (**E**). Vertical lines represent rough thresholds for statistical significance (as in **A**). The unity line (y=x) is shown. (**F**) Example evoked response potentials for a shared electrode in Bravo-3. Error bars represent standard error of the mean. (**G**) Mean HGA, theta power, and beta power during reading, listening, and attempted speech across electrodes with significant task modulation (as in **A**) for listening, reading, and attempted speech (** P < 0.01, **** P < 0.0001; two-sided Wilcoxon rank-sum tests with 9-way Holm-Bonferroni correction). (**H-J**) Bravo-3 results (top) and Bravo-1 results (bottom). (**H**) Shown is classification accuracy for the speech-verification model, trained on specific regions and feature streams in Bravo-3. Distributions are over 10 non-overlapping cross-validation folds (*** P < 0.001, ** P < 0.01; Two-sided Wilcoxon rank-sum tests with 6-way Holm-Bonferroni correction). Statistical results are shown for adjacent distributions after sorting by median classification accuracy. (**I**) Speech-verification model electrode contributions (all, HGA + LFS) in Bravo-3. (**J**) Confusion matrix for predictions from the full speech-verification model in Bravo-3. In (**A,I,L**) the black region outline indicates the midPrCG and blue shading the precentral gyrus. **(A,I)** MidPrCG outlined in black with the PrCG shaded blue.

We next asked if shared electrodes had different spectrotemporal responses for each function. We found that, at shared electrodes, attempted speech generally evoked the highest maximum HGA followed by reading and listening (Fig. 2F, Fig. S5). Shared electrodes also had increased HGA before the go-cue, in contrast to reading and listening (Fig. 2F, Fig. S5), and further differed in their low-frequency content; attempted speech had significantly stronger suppression in theta- (5-12 Hz) and beta-band (20-30 Hz) power relative to reading and listening (Fig. 2G; two-sided Wilcoxon rank-sum tests with 12-way Holm-Bonferroni correction). This aligns with literature showing that there is low-frequency desynchronization during intact, healthy movements [43,44] and synchronized low-frequency signals during perceived speech [45,46]. Thus, despite shared electrodes being responsive across functions, spectrotemporal response profiles are distinct.

We next investigated the contribution of these broad anatomical and spectral differences to differentiating between reading, listening, and attempted speech. Offline, we limited training of the speech-verification model to each different anatomical region, one at a time. In Bravo-3, discriminability is strong across anatomical areas, with the best performance from the precentral gyrus (Fig. 2H; two-sided Wilcoxon rank-sum tests with 12-way Holm-Bonferroni correction). In line with our spectral analysis (Fig. 2G), incorporation of low frequency signals (LFS) further improved performance over HGA alone (Fig. 2H, top). Interestingly, electrodes that contributed most to performance are in the midPrCG and STG, providing further evidence that, despite shared activations, underlying spectrotemporal profiles are differentiable across tasks (Fig. 2I, top). In Bravo-1, performance was also highest using electrodes from the precentral gyrus and adding LFS increased performance (Fig. 2H, bottom; two-sided Wilcoxon rank-sum tests with Holm-Bonferroni correction for multiple comparisons). Similar to Bravo-3, electrodes driving performance were in the midPrCG (Fig. 2I, bottom). Although overall accuracy was lower in Bravo-1, this was due to confusability between listening and reading with strong separation of attempted speech from reading or listening (Fig. 2J, bottom).

During daily use of a speech neuroprosthesis, non-speech movements (i.e. hand, arm, leg, eye, or head movements; Fig. 2B) also have the potential to falsely engage the system and activate the SMC [47]. While we did not collect online non-speech movement data with use of the full system (as in Fig. 1), we tested this scenario by simulating how well the speech-verification model could distinguish between attempted-speech, listening, reading, and attempted non-speech movements (see Methods “Isolated-target task”). Given that electrode contributions of the speech-verification and detection models were significantly correlated (Fig. S6), we performed this analysis on the speech-verification model. We found that the speech-verification model retained high performance, and similar anatomical trends, when adding the non-speech movement class, though the performance was minimally reduced (82.6% versus 86.0% for Bravo-1 and 97.1% versus 98.3% for Bravo-3; P < 0.01, two-sided Wilcoxon rank-sum test; Fig. S7).

### Content selectivity of shared activations across attempted speech and perception

Our prior analyses focused on whether cortical activity across attempting speech, listening, or reading was discriminable. We next investigated whether models could be trained to decode activations within each task into the corresponding word in the 10-word vocabulary. As a secondary question, we asked if these models could generalize across tasks, which would indicate shared electrodes represent content in the same manner across tasks. We trained distinct classifiers on cortical-activity from each anatomical region during attempted-speech, reading, and listening, using the 10-word vocabulary (Fig. 2A). While electrodes were activated by reading (Fig. 2A), reading content was not decodable above chance from any anatomical region covered by the participants’ ECoG arrays (two-sided Wilcoxon signed-rank test against chance level of 10% with 12-way Holm-Bonferroni correction). Listening was only decodable from the temporal lobe in Bravo-3. Attempted speech was decodable across all regions, including the temporal lobe (Fig. 3A). We next asked if the temporal lobe represented words in the same manner during listening and attempted speech, given that words could be decoded in multiple tasks. We used 10-word classifiers trained on attempted speech to predict on listening trials and vice versa, finding low generalizability of the models across functions (Fig. 3B). Additionally, electrode contributions for decoding attempted speech versus for decoding listening were not significantly correlated (P = 0.74, Spearman correlation permutation test; Fig. 3C). Together, these results suggest that while neural populations have shared activations across attempted speech and listening, they do not leverage identical representations for each function. Practically, while reading and listening may generate false positives without proper system design (Fig. 1D,G), it is unlikely that an ECoG-based attempted-speech model will accurately decode the content of what the user is listening to or reading.

**Figure 3.**
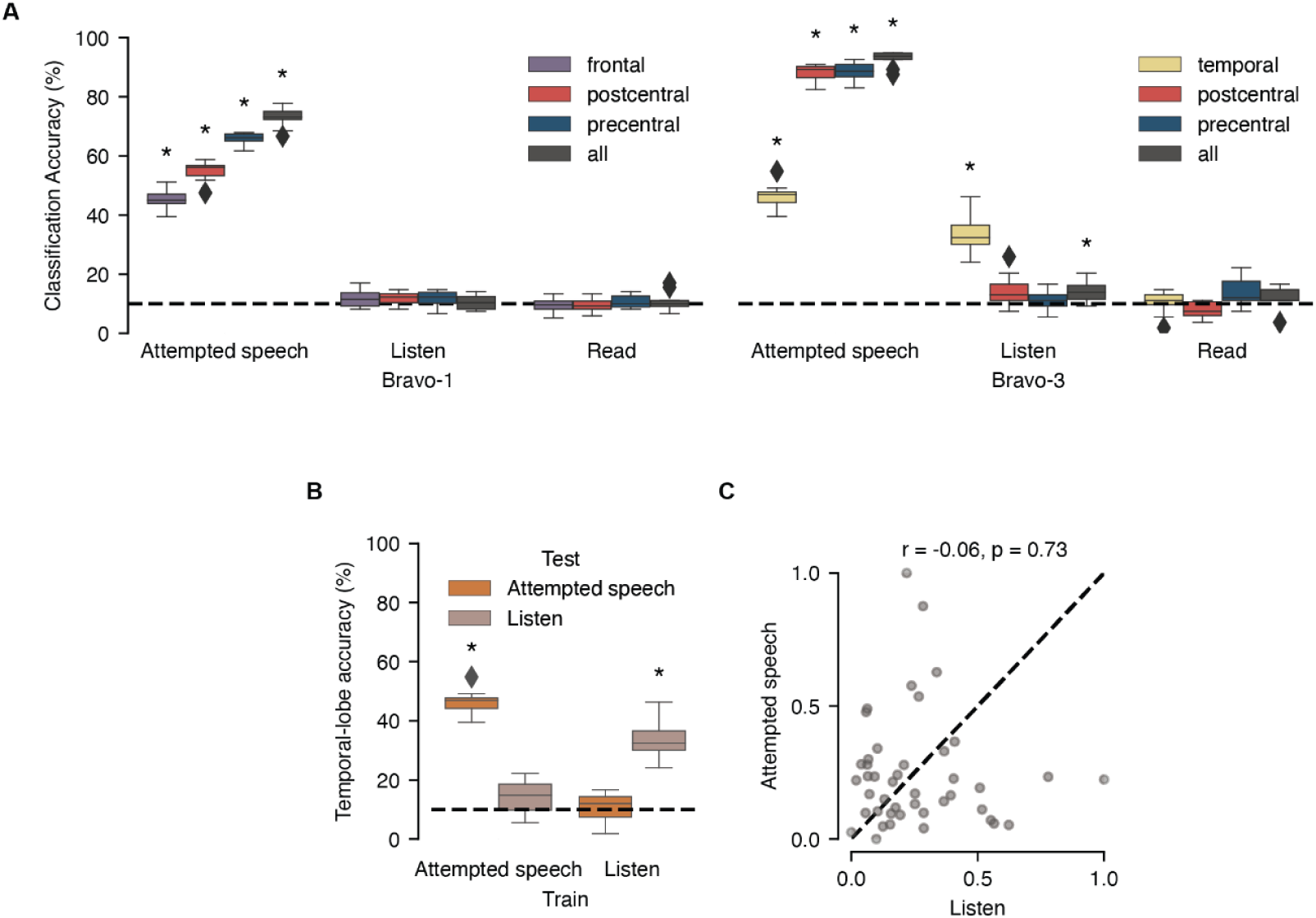
Distinct representations for attempted speech, listening, and reading on speech cortex . (**A**) Shown is classification accuracy by anatomical region on the isolated-word set during reading, listening, and attempted speech. Statistical annotations reflect above-chance performance (*P < 0.05; two-sided Wilcoxon signed-rank test against median chance level of 10% with 12-way Holm-Bonferroni correction for multiple comparisons). (**B**) For the Bravo-3 temporal lobe attempted speech and listening classification models, accuracy is shown for evaluating models across tasks. Statistical annotations reflect above-chance performance (*P < 0.05; two-sided Wilcoxon signed-rank test against median chance level of 10% with 4-way Holm-Bonferroni correction for multiple comparisons). (**A-B**) distributions are over 10 non-overlapping cross validation folds of the data. (**C**) Scatterplot of electrode contributions across temporal lobe electrodes for attempted speech and listening models (r = -0.06 and P = 0.72; Spearman correlation and permutation test).

### A role for shared electrodes in motor planning

Despite the fact that reading activations were not word-selective (Fig. 3A), we assessed whether they were stronger for linguistic content, which would suggest a meaningful role in speech processing rather than reflecting any transient change in visual input. This is a particularly important question, given the proximity of the midPrCG to the frontal eye fields [39] and visual attention network, which transiently activates in response to abrupt changes in the visual field (such as text appearing on the screen) [48,49]. To test this, we presented the participants with sentences and false fonts, such as “δƱΦϞΨƟ” [30], to read, in addition to the isolated words. False fonts have been used in prior functional MRI work to contrast transient visual versus speech processing [30,50,51]. Across both participants, evoked HGA was stronger for reading the isolated words versus false fonts (P < 0.001, two-sided Wilcoxon signed-rank test; Fig. 4A,B). The few electrodes with comparable evoked HGA were located near to the frontal eye fields [39] in Bravo-3 (Fig. 4A, Fig. S8). We also presented the participant with sentences that stayed on the screen for the same time as words. Sentences evoked sustained activations across the 0 to 2 s analysis window (Fig. 4A, C), suggesting active processing of the stimuli rather than transient activation related to a visual change on the screen.

**Figure 4.**
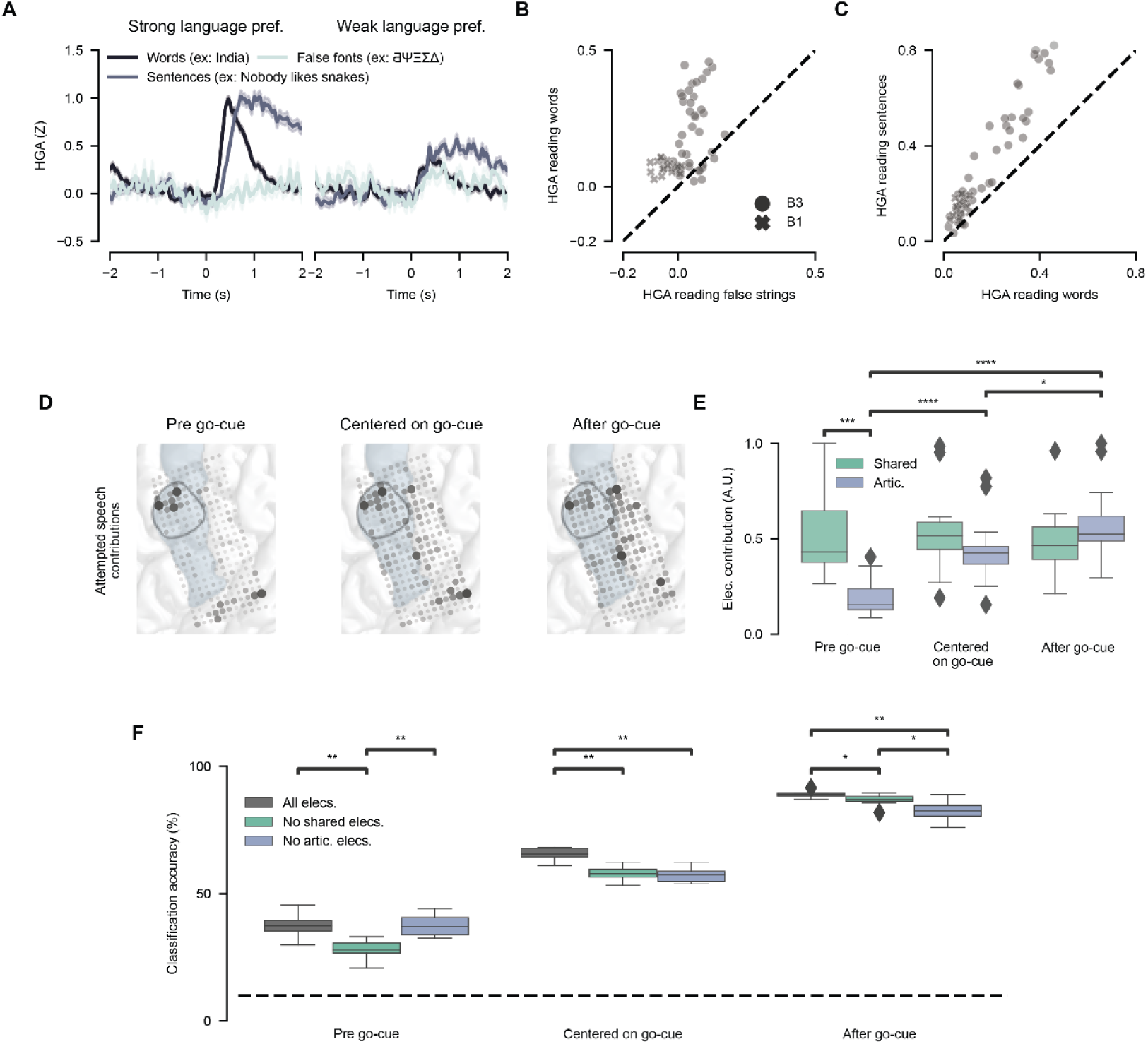
A role for shared electrodes in speech-motor planning. (**A**) Shown are example evoked response potentials (ERPs) from two electrodes in Bravo-3 that are task-modulated during reading (as in Fig. 2A). ERPs are aligned to the onset of reading words from the isolated-word set, sentences, and false fonts. (**B**) For each reading-responsive electrode, the average HGA (0-2s) is scattered between reading words from the isolated-word set and false fonts. (**C**) For each reading-responsive electrode, the average HGA (0-2s) is scattered between reading words from the isolated-word set and sentences. (**D**) Shown are electrode contributions for attempted-speech models, trained on different windows of time around the go-cue: strictly before the go-cue, centered on the go-cue, and strictly after the go-cue. The black region outline indicates the midPrCG and the blue shading the precentral gyrus. (**E**) The average electrode contribution for articulatory encoding and shared electrodes in each decoding window is quantified (* P < 0.05, *** P < 0.001, **** P < 0.0001; two-sided Wilcoxon rank-sum tests with 9-way Holm-Bonferroni correction for multiple comparisons). Error bars reflect standard error of the mean. (**F**) Accuracy of the classification model across the three decoding windows and three model conditions. Performance was measured with 10-fold cross validation (* P < 0.05, ** P < 0.01; two-sided Wilcoxon rank-sum tests with 9-way Holm-Bonferroni correction for multiple comparisons).

Shared neural populations that are activated across movement and audiovisual processing have been proposed to play a role in motor planning of coordinated movements [15,23]. We hypothesized that shared electrodes have a specific role in speech-motor planning given multiple observations in our data. First, shared electrodes in Bravo-3 had multisensory properties and reading activations reflected speech processing. Second, shared electrodes had HGA above baseline before speech attempts (a requirement of motor planning activity) [52]. Finally, shared electrodes were different from those electrodes which encoded specific articulator movements (Fig. 2B) [15,24]. To investigate this hypothesis, we trained classifiers on attempted-speech data from three sequential windows around the go-cue: one exclusively before the go-cue, one centered on the go-cue, and another exclusively after the go-cue. We found that shared electrodes in the midPrCG were most important for decoding attempted speech before the go-cue whereas articulatory encoding electrodes, around the central sulcus, were most important for decoding during the window after the go-cue (Fig. 4D,E; Fig. S9). Notably, shared electrodes not only contributed to decoding before the go-cue, but their contributions remained consistent across windows, in contrast to articulatory encoding electrodes (Fig. 4E). To further support this result, we compared accuracy at each window under three conditions: “all electrodes”, “no shared electrodes” (all 253 electrodes minus the 42 shared electrodes), and “no articulatory electrodes” (all 253 electrodes minus 41 articulatory encoding electrodes). We found that performance before the go-cue was lowest when shared electrodes were removed whereas performance after the go-cue was lowest when articulatory electrodes were removed (Fig. 4F). This further supports shared activity in the midPrCG has a distinct role in motor-planning before attempted speech (Fig. 4E).

## Discussion

We analyzed the impact of shared perceptual and motor representations in the SMC on speech neuroprostheses in two participants with vocal-tract paralysis. When speech detection and verification models were trained on cortical activity during reading and listening, there were zero false-positive activations of the system during sustained periods of listening, reading, or mental imagery tasks. Furthermore, decoding performance was not disrupted while listening to audio during speech attempts. Offline, we found that the SMC contained neural populations with shared representations for attempted speech, reading, and listening, but the spectrotemporal properties of these activation patterns were strongly discriminable, which supported the system differentiating between perceptual tasks and attempted speech.

A commonly cited concern, from both potential users and experts in the field, is that speech neuroprostheses will be activated by and decode internal thoughts or visual/auditory perception that are not intended to be communicated [53–57]. Indeed, one study found that listening falsely engaged a speech decoder, resulting in unintended output from the system [27]. It is important that end-users of neuroprostheses retain volitional control over the system to encourage long-term safety, efficacy, and adoption of the technology [25,55]. We found that with proper design, decoders can maintain complete specificity to intended-speech during perceptual tasks. We also demonstrated that our speech-decoding system was not falsely activated by non-speech mental imagery and, offline, demonstrated that non-speech movements were highly discriminable from speech attempts. An important future direction is to extend these results to real-world and clinical scenarios, considering additional sources of interference such as combined audiovisual stimuli [58] and non-speech orofacial gestures [59,60], such as facial expressions.

Although the midPrCG had the most shared electrodes, it was also a critical region for discriminating perceptual functions from attempted speech. Our results suggest that this was due to the differences in the spectrotemporal response patterns for each task, including activations before the onset of speech attempts. Though reading and listening content was not strongly decodable from the midPrCG, the midPrCG was important for decoding attempted speech before the attempt. These findings corroborate other studies that associate shared neural populations with planning coordinated movements [15,16,23,24]. The spatial localization of shared activity aligns with the hypothesis that the midPrCG is an area involved in speech-motor planning distinct from the direct execution of articulation [24,61]. This functional distinction also highlights a spatial distinction–neural populations tuned to intended movements of specific articulator groups localized along the central sulcus in the vSMC while neural populations with shared activity, important for speech-motor planning, localized to the midPrCG. Further understanding of differences between the vSMC and midPrCG, along with developing algorithms to harness the unique information from each, has the potential to improve speech decoding accuracy.

It is important to note that our study is limited by a sample size of two participants with a similar etiology of speech loss, due to the rare nature of recording such data. Future studies should better sample the space of end-users, including people who are fully locked-in [62]. Our two participants also had slightly different ECoG array coverage, with Bravo-3 having a larger, denser array that was centered on the central sulcus and extended ventrally into the temporal lobe. While our results align with studies in healthy speakers that also find shared representations in the SMC [19,63], more studies across many people with different types of paralysis are needed to confirm that our findings generalize across potential users, including those who are fully locked-in. Further, comparisons to healthy speakers are needed to holistically assess electrophysiological changes after paralysis. Additionally, we tested our speech decoding with a restricted vocabulary of words; though a similar framework may scale to larger vocabulary approaches [3,4,6]. In cases where lower latency decoding is preferred, the low latency speech detection model could be used without the longer latency speech-verification model. Alternatively, a single holistic model could be used to perform speech detection and decoding, such as the case in modern automatic speech recognition algorithms [64]. Nonetheless, future studies are needed to reinforce the findings of this work in the context of larger-vocabulary decoding and independent-use settings.

To create reliable neuroprostheses that are adopted by people with paralysis, it is important to develop high-specificity decoding frameworks that maintain performance during common daily functions that activate the SMC. In this study, we demonstrate a speech decoding system that meets these goals. Further, we found that shared neural populations on the SMC localized primarily to the midPrCG and may play a specific role in speech-motor planning that could be leveraged by future neuroprostheses.

## Data Availability

All data needed to evaluate the conclusions in the paper are present in the paper and/or the
Supplementary Materials. Summary data and code to recreate the main results of the
manuscript will be provided in a github repository and made public upon publication.

## Acknowledgements

We thank our participants Bravo-1 and Bravo-3 for their unwavering dedication and commitment to the project. We thank members of the Chang Lab for feedback and Viv Her for administrative support. We also want to thank our participant’s families and caregivers for logistic support. Ben Speidel helped with imaging reconstruction of the patient’s pial surface and electrode array and Deborah Levy helped by providing stimuli to investigate false fonts.

## Funding

This work was supported by the National Institutes of Health (grant NINDS 5U01DC018671), Joan and Sandy Weill Foundation, Susan and Bill Oberndorf, Ron Conway, Graham and Christina Spencer, and William K. Bowes, Jr. Foundation supported authors A.B.S, J.R.L, V.R.A, C.M.K, I.P.H, K.T.L, S.B, D.A.M, and E.F.C. K.T.L is also supported by Rose Hills and Noyce Foundations as well as the National Science Foundation GRFP. A.B.S is supported by the National Institute On Deafness And Other Communication Disorders of the National Institutes of Health award number F30DC021872. Authors A.T.C. and K.G. did not have relevant funding for this work.

## Author contributions

A.B.S and J.R.L conceptualized and led the project. A.B.S developed the classification and speech-verification models and performed neural population analyses. J.R.L. developed speech detection models. A.B.S, J.R.L, V.R.A., and C.M.K led data collection with help from K.T.L, S.B, I.P.H, and D.A.M. A.B.S and J.R.L designed the tasks which were implemented by D.A.M and I.P.H. A.B.S and J.R.L wrote the manuscript and generated the figures with input from the other authors. A.T.C., K.G., and E.F.C. performed regulatory and clinical supervision. E.F.C. and K.G. conceived and supervised the clinical trial.

## Competing interests

D.A.M., J.R.L. and E.F.C. are inventors on a pending provisional UCSF patent application that is relevant to the neural-decoding approaches surveyed in this work. E.F.C. is an inventor on patent application PCT/US2020/028926, D.A.M. and E.F.C. are inventors on patent application PCT/US2020/043706 and E.F.C. is an inventor on patent US9905239B2, which are broadly relevant to the neural-decoding approaches surveyed in this work. EFC is co-founder of, and DAM is a director at, Echo Neurotechnologies, LLC. All other authors declare no competing interests.

## Data and Materials availability

All data needed to evaluate the conclusions in the paper are present in the paper and/or the Supplementary Materials. Summary data and code to recreate the main results of the manuscript will be provided in a github repository and made public upon publication.

## Notes

### Clinical Trial

NCT03698149

### Author Declarations

This study was conducted as part of the BCI Restoration of Arm and Voice (BRAVO) clinical trial (ClinicalTrials.gov; NCT03698149) approved by the Federal Drug Association, University of California San Francisco IRB, and the National Institutes of Health, with all ethical regulations rigorously followed in accordance with the Declaration of Helsinki. Although this data was collected as a part of this clinical trial, the data presented in this manuscript is not to inform concrete conclusions regarding the primary outcomes of the trial.

