## Supplement for "Implications of shared motor and perceptual activations on the sensorimotor cortex for neuroprosthetic decoding"

Figure S1. Artifact that affected 3 trials in the online evaluation blocks. 2
Figure S2. Correlation of listening activity with audio volume. 3
Figure S3. Effect of speech decoding system ablations on false positive and negative performance. 4
Figure S4. Simulations to assess speech-verification model performance 6
Figure S5. Temporal dynamics of shared electrodes during attempted speech, listening, and reading. 7
Figure S6. Significant correlation between electrode contributions to the speech detection and speech-verification models. 8
Figure S7. Separability of non-speech movements from attempted-speech, reading, and listening. 9
Figure S8. Electrodes with comparable HGA for reading words and false fonts localize around the frontal eye fields. 10
Figure S9. Anatomical characteristics of electrodes important for decoding attempted speech pre go-cue and after the go-cue. 11
Table S1. 10-word utterance sets for each participant 12
Table S2. Parameters corresponding to the speech detection model. 13
Table S3. Parameters corresponding to the speech-verification model. 14
Table S4. Parameters corresponding to the speech-classification model. 15
Table S5. Participant survey on preference for latency versus specificity in different daily-use scenarios. 16
Video S1. Speech decoding alternating with reading in Bravo-3 17
Video S2. Speech decoding during the listening condition in Bravo-1 18
Video S3. System specificity for attempted speech during listening in Bravo-1 19

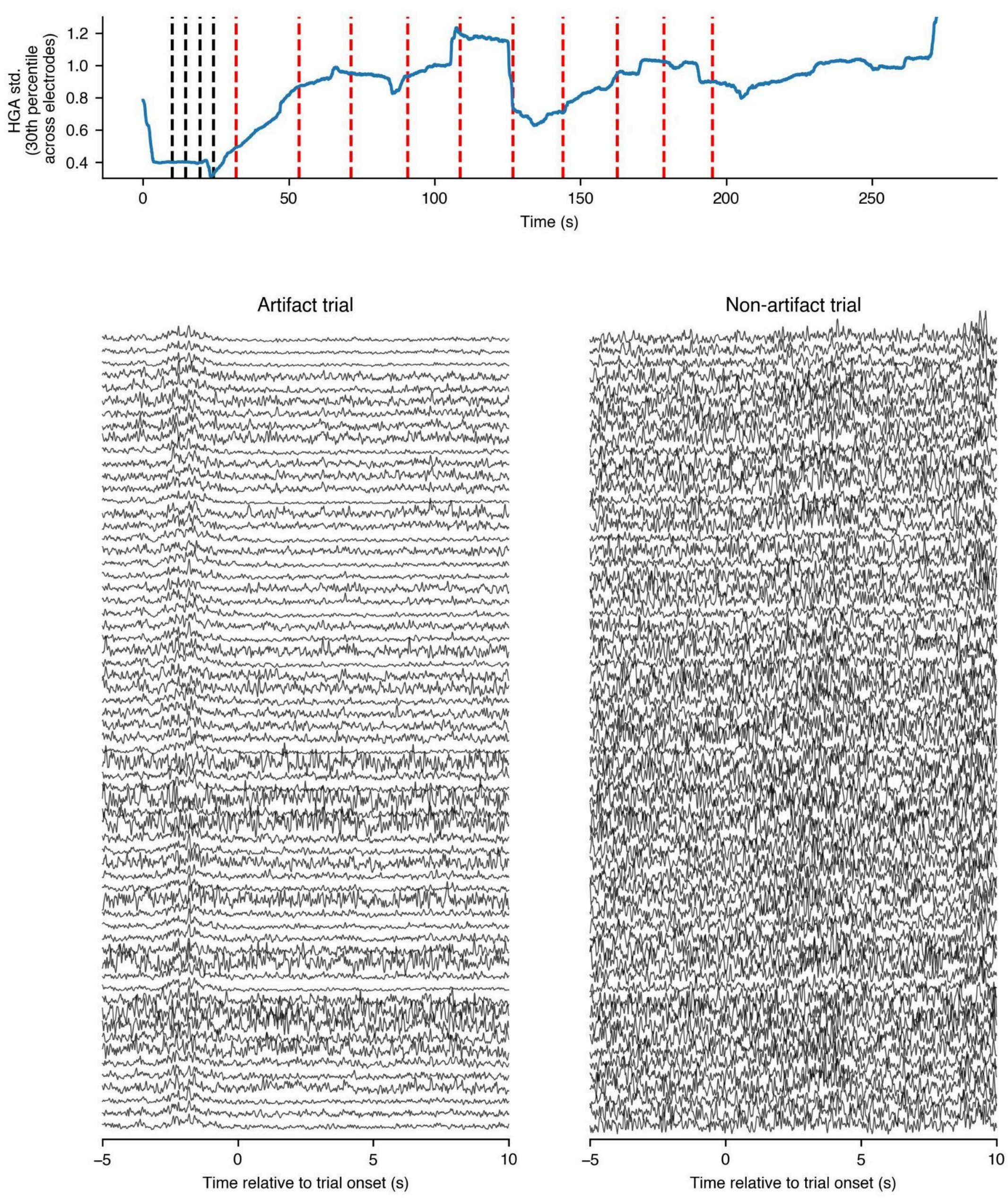


**Figure S1. Artifact that affected 3 trials in the online evaluation blocks.**

Shown is a sample artifact that occurred during three trials of online evaluation with Bravo-1. The standard deviation of the high-gamma amplitude is visualized over time **(top)**. This metric was computed by first taking the HGA standard deviation for each electrode over the window [-5, 10] relative to each timepoint. Next, to quantify whether a proportion of electrodes' had decreased their variance, the 30th percentile of the HGA std across electrodes at each timepoint was taken. Sample trials with and without an artifact **(bottom).**

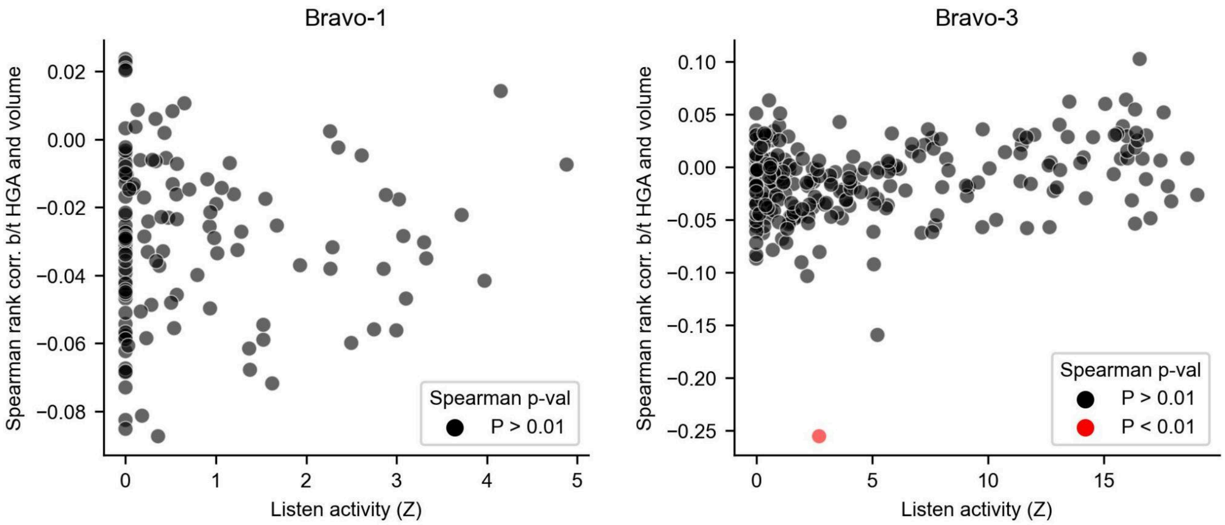


**Figure S2. Correlation of listening activity with audio volume.**

For Bravo-1 and Bravo-3, the listening activity (as in Fig. 2A) is plotted along with the spearman correlation between that electrode's HGA and volume, across listening blocks. There is not a strong relationship between Listen activity and the Spearman correlation between that electrode's HGA and audio volume. Overall, only one electrode had a significant correlation ($P < 0.01$, Spearman correlation with Holm-Bonferroni correction across electrodes within each participant).

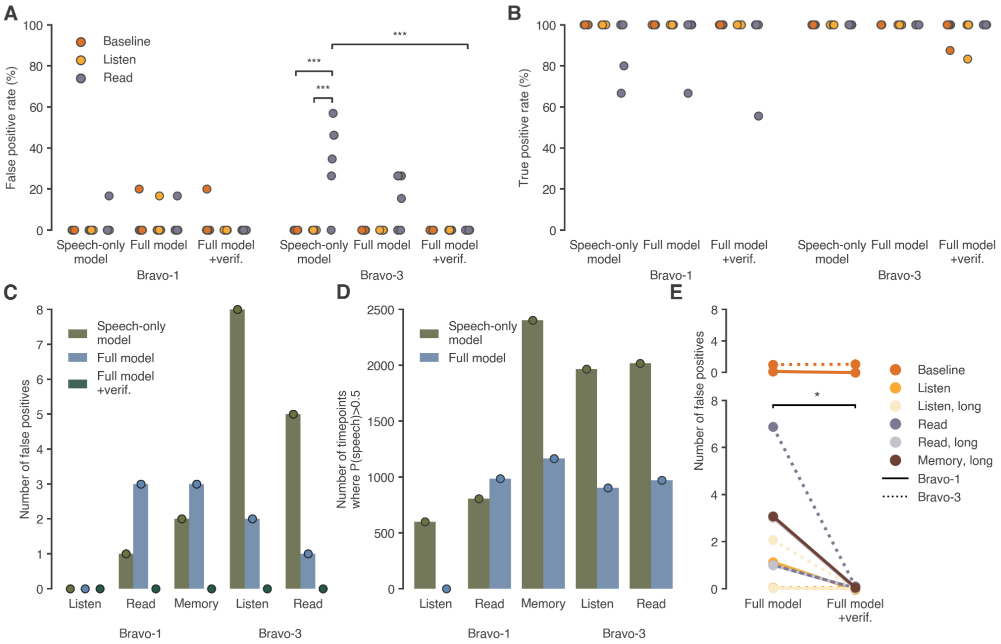


**Figure S3. Effect of speech decoding system ablations on false positive and negative performance.**

**A.** The false positive rate (FPR) of the system with only a speech-detection model trained only on attempted speech (no reading or listening data), the full model (trained with reading and listening data but without the speech-verification classifier), and the full speech decoding system during baseline, listening, and reading blocks. FPR generally increases as system protections are removed (i.e. not using the speech-verification model and not giving the speech-detection model examples of listening and reading to dissociate from attempted speech), however, they are only significantly increased from baseline during the reading condition when using the speech-only model with Bravo-3 and when comparing the speech-only model to the full system during the reading condition with Bravo-3 (*** $P < 0.0001$, Fisher exact test with 18-way Holm-Bonferroni correction). **B.** The true positive rate (TPR) for each of the system ablations in **A** during baseline, listening, and reading blocks. TPR is not significantly different for any of the conditions or system ablations, for either participant. Panels **A** and **B** are extensions of panels **D** and **E** in main text Figure 1. **C.** The absolute number of false positives for each of the system ablations during “longform” blocks of listening and reading, where there were no speech attempts and only the perceptual content. For Bravo-1, this also included memory reflection during an autobiographical interview block (see Methods). For Bravo-1, who had much less of a pronounced overlap between reading, listening, and attempted speaking neural responses, a speech-only model outperformed a model that included examples of listening. Using the full system still resulted in the best performance. For Bravo-3, including reading and listening in the speech-detection model and then adding the speech-verification model both decreased the number of false positives. **D.** The number of time points (at 200 Hz) that had a speech probability greater than the probability threshold of 0.5 for each of the system ablations and “longform” blocks. These time points are not all consecutive and can reflect brief spikes in the probability that do not result in false positive events. For reference, 2500 time points corresponds to a cumulative 12.5 seconds of time points with a speech probability above 0.5, while 500 corresponds to 2.5 seconds. Even though including reading and listening in the speech-detection model (“Speech-only model” versus “Full model”) did not have an effect or resulted in slightly more false positive events (as shown in **C**, with no effect on listening, 2 more events during reading and 1 more event during memory reflection), the full model resulted in few time points with high speech probability. This suggests that while it was not enough to decrease the number of events, the addition of reading and listening examples still generally improves the accuracy of the continuously predicted probabilities. **E.** The number of false positives during non-baseline conditions significantly decreased with the addition of the speech-verification model (* P < 0.01, one-sided Wilcoxon signed-rank test). Lines plotted for both regular and “long” versions of the non-baseline conditions during which listening or reading was present. Solid lines indicate Bravo-1 while dotted lines indicate Bravo-3.

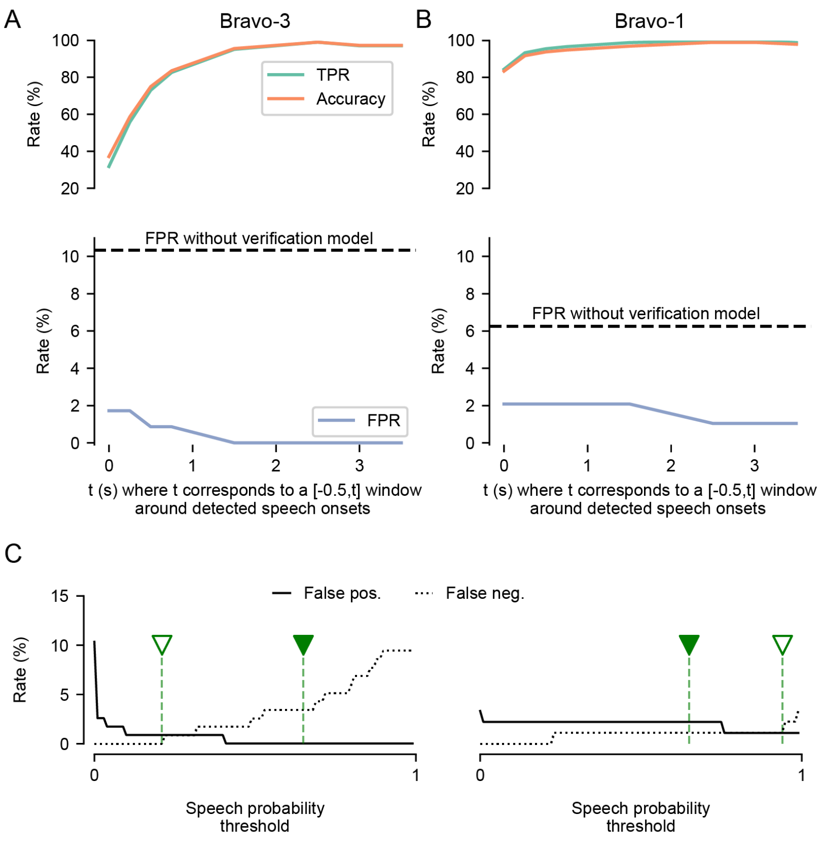


**Figure S4. Simulations to assess speech-verification model performance**

**A**. For Bravo-3, the accuracy of the speech-verification model in correctly accepting/rejecting detected speech-events using various time windows is shown. An accuracy of 100% means that all detected speech events corresponding to true attempts are accepted and all detected speech events not corresponding to true speech attempts are rejected. Accuracy is plotted as a function of context length of neural features passed to the verification model after detected onset. Accuracy and the true positive rate (top) are shown using time windows spanning [-0.5, t] around a detected event, where t is iteratively increased to 3.5 s. The online false positive rate (bottom; as in Fig. 1D) is also shown as a function of context length, with the horizontal dashed line marking the false positive rate without use of the verification model. **B.** The same as (**A**) for Bravo-1. **C.** The speech-verification model predicts how likely a detected event is attempted speech, reading, or listening, producing a probability distribution over these 3 classes that sums to 1. The attempted-speech probability threshold, over which to accept a detected event, may be adjusted. Here, we plot the false positive and negative rates as a function of the probability threshold used to accept/reject detected events. Vertical green lines with filled markers note the threshold used during real-time testing (0.65) while unfilled markers note the optimal threshold that minimizes both error rates (offline). As expected, lower thresholds are more generous in accepting events and therefore result in a low FNR but increased FPR, and vice versa for higher thresholds. This parameter is customizable and allows participants to trade-off between FPR and FNR.

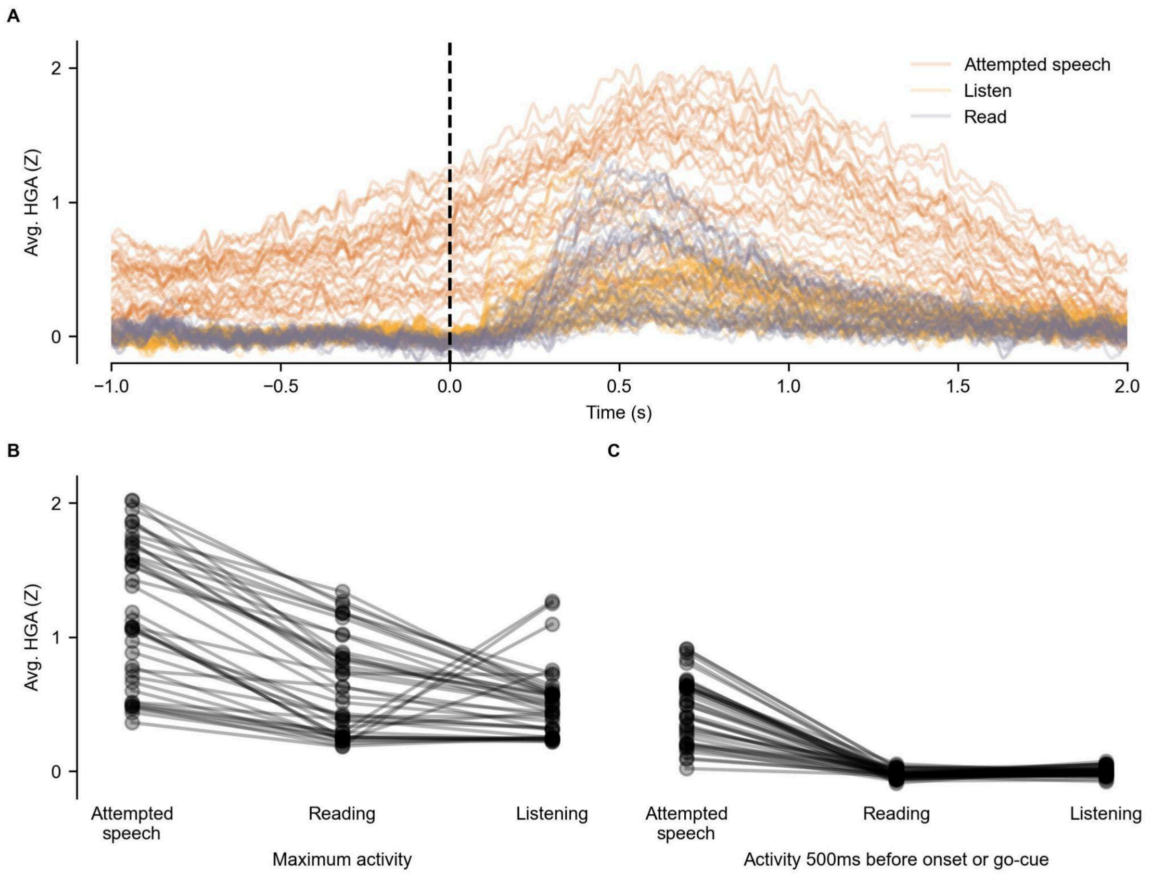


**Figure S5. Temporal dynamics of shared electrodes during attempted speech, listening, and reading.**

**A.** Shown are mean evoked response potentials (ERPs) for attempted-speech, listening, and reading. ERPs are shown across all shared electrodes. **B.** For each shared electrode, the maximum HGA evoked by attempted speech, reading, and listening is shown. **C.** For each shared electrode, the evoked HGA 500ms before the onset/go-cue for attempted speech, reading, and listening is shown.

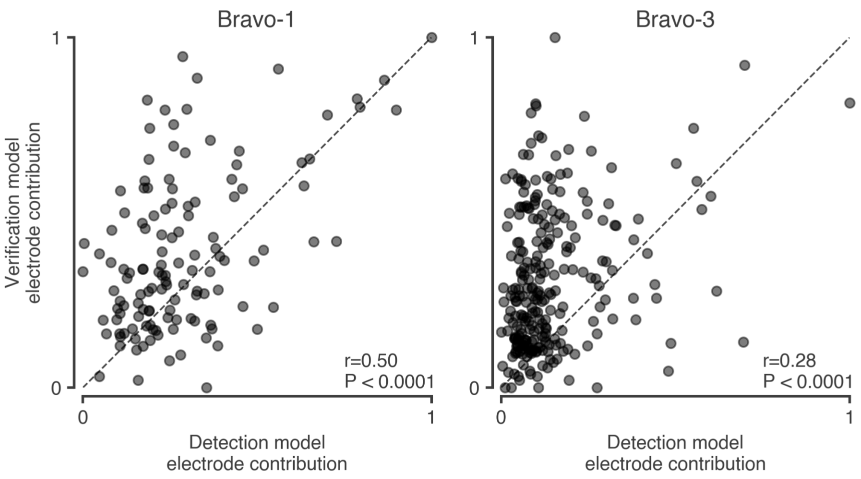


**Figure S6. Significant correlation between electrode contributions to the speech detection and speech-verification models.**

Scatterplot of electrode contributions from the speech detection and speech-verification models for Bravo-1 (left) and Bravo-3 (right). For both participants, contributions are significantly correlated (P < 0.0001; Non-parametric Spearman correlation and permutation test), especially for the strongest contributing electrodes.

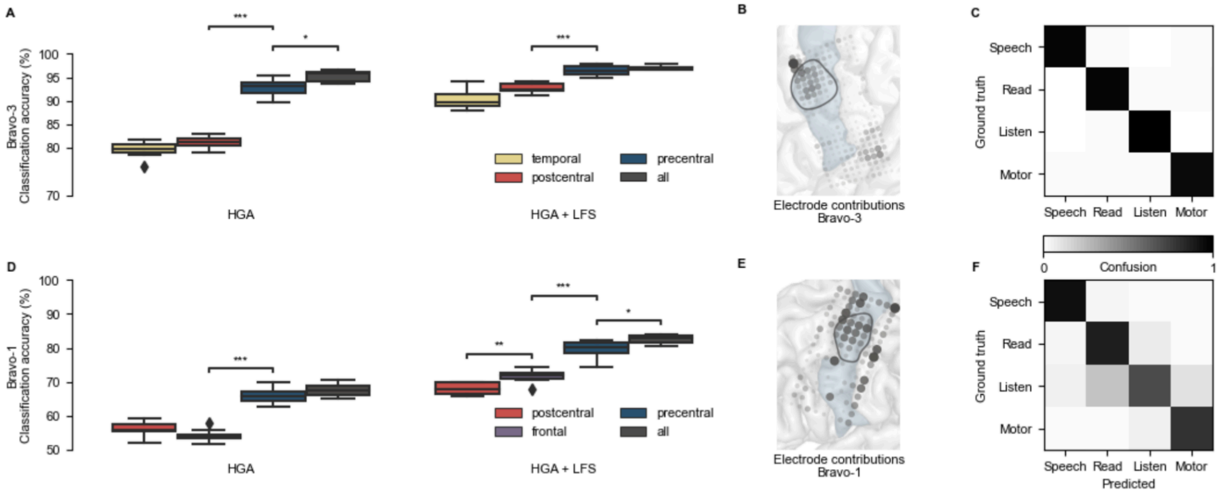


**Figure S7. Separability of non-speech movements from attempted-speech, reading, and listening.**

Extension of Figure 2H-M to add an additional class of non-speech movements. **a,** Shown is classification accuracy for the speech-verification model, trained on specific regions and feature streams in Bravo-3. Distributions are over 10 non-overlapping cross validation folds (* $P < 0.05$, ** $P < 0.01$, ***$P < 0.001$; Two-sided Wilcoxon rank-sum tests with 6-way Holm-Bonferroni correction for multiple comparisons). Statistical results are shown for adjacent distributions after sorting by median classification accuracy. **b** Shown are electrode contributions to the full model (all, HGA + LFS) in Bravo-3. **c,** Shown is a confusion matrix for predictions from the full model in Bravo-3. **d-f,** The same as (**a-c**) for Bravo-1. In (**b,e)** the black region outline indicates the midPrCG and blue shading the precentral gyrus.

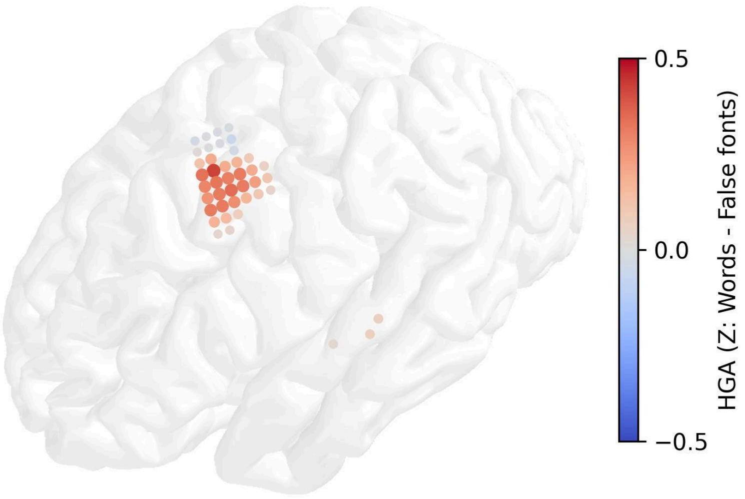


**Figure S8. Electrodes with comparable HGA for reading words and false fonts localize around the frontal eye fields.**

For reading-modulated electrodes, the difference in mean HGA during the first 2 seconds of reading real words vs false fonts is shown.

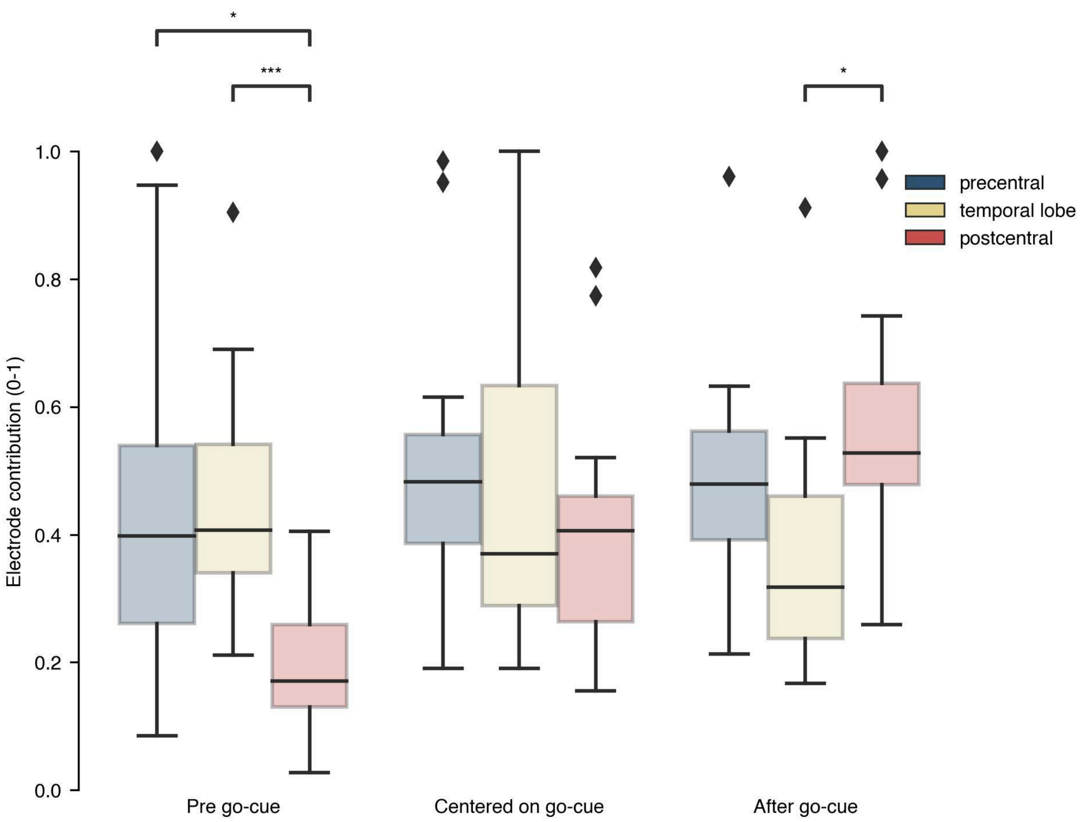


**Figure S9. Anatomical characteristics of electrodes important for decoding attempted speech pre go-cue and after the go-cue.**

Shown are electrodes contributions by anatomical regions for each decoding model trained on different windows around the go-cue. To be considered in the analysis, an electrode must be within the top 10th percentile of contributing electrodes for at least one of the decoding windows (* $P < 0.05$, *** $P < 0.001$; two-sided Wilcoxon rank-sum tests with Holm-Bonferroni correction).

**Table S1. 10-word utterance sets for each participant**

| Participant | Utterances | |
|---|---|---|
| Bravo-1 | Pancho | Despierto |
| | Alimento | Vehicle |
| | Enfermero | Speak |
| | Ropa | Funny |
| | Comer | Awake |
| Bravo-3 | X-ray | India |
| | Bravo | Yankee |
| | Kilo | Sierra |
| | Foxtrot | Charlie |
| | Whiskey | Hotel |

**Table S2. Parameters corresponding to the speech detection model.**

| Participant | Parameter | Value |
|---|---|---|
| Bravo-1 | Number of LSTM layers | 3 |
| | Nodes per LSTM layer | 128, 128, 32 |
| | Dropout rate | 0.5 |
| | Optimizer | Adam |
| | Learning rate (reduced on plateau) | 0.01 |
| | Smoothing window size | 100 time points |
| | Probability threshold | 0.5 |
| | Time threshold | 100 time points |
| Bravo-3 | Number of LSTM layers | 3 |
| | Nodes per LSTM layer | 128, 96, 32 |
| | Dropout rate | 0.5 |
| | Optimizer | Adam |
| | Learning rate (reduced on plateau) | 0.001 |
| | Smoothing window size | 100 time points |
| | Probability threshold | 0.5 |
| | Time threshold | 100 time points |

**Table S3. Parameters corresponding to the speech-verification model.**

| Participant | Parameter | Value |
|---|---|---|
| Bravo-1 | Number of GRU layers | 2 |
| | Nodes per GRU layer | 220 |
| | Dropout rate | 0.7 |
| | CNN Kernel size and stride | 8 |
| | Time window | [-0.5,3.5] |
| | weight decay | 0.0001 |
| | Optimizer | Adam |
| | Learning rate (reduced on plateau) | 0.0005 |
| Bravo-3 | Number of GRU layers | 2 |
| | Nodes per GRU layer | 220 |
| | Dropout rate | 0.7 |
| | CNN Kernel size and stride | 8 |
| | Time window | [-1,3] |
| | weight decay | 0.0001 |
| | Optimizer | Adam |
| | Learning rate (reduced on plateau) | 0.0005 |

**Table S4. Parameters corresponding to the speech-classification model.**

| Participant | Parameter | Value |
|---|---|---|
| Bravo-1 | Number of GRU layers | 3 |
| | Nodes per GRU layer | 260 |
| | Dropout rate | 0.8 |
| | CNN Kernel size and stride | 4 |
| | Time window | [-0.5,3.5] |
| | weight decay | 0.0001 |
| | Optimizer | Adam |
| | Learning rate (reduced on plateau) | 0.0001 |
| Bravo-3 | Number of GRU layers | 2 |
| | Nodes per GRU layer | 274 |
| | Dropout rate | 0.54 |
| | CNN Kernel size and stride | 4 |
| | Time window | [-1,3] |
| | weight decay | 0.0001 |
| | Optimizer | Adam |
| | Learning rate (reduced on plateau) | 0.0005 |

**Table S5. Participant survey on preference for latency versus specificity in different daily-use scenarios.**

We provided our participants with the following survey to assess their preference for latency versus specificity of a future speech-neuroprosthetic system in various daily-use scenarios: “For each of the following daily-use settings, please rate how important latency versus specificity of the system would be to you. You can use the scale below, which ranges from 0-10 where 0 indicates a preference for near instantaneous latency but some false positives, 5 indicates no preference, and 10 indicates a preference for no false positives but 1-2 s latency.”

| **Situation** | **Participant** | |
|---|---|---|
| | **Bravo-1** | **Bravo-3** |
| Informal conversation with friends | 8 | 1 |
| Informal conversation with family | 9 | 1 |
| Conversation with friends about an important topic | 9 | 9 |
| Conversation with family about an important topic | 10 | 9 |
| Formal, work presentation | 8 | 9 |
| Doctor’s appointment | 8 | 9 |
| Legal proceedings (e.g. jury, witness, filing a report) | 10 | 9 |
| Communication with caregiver for a personal need (i.e. that you are thirsty or hungry) | 8 | 1 |
| Communication with caregiver for a medical need (i.e. that you need your medication now) | 8 | 1 |
| Ordering coffee or food at a restaurant | 8 | 3 |
| Dictating an email, text, or writing of some sort using a text editor or writing software | 6 | 9 |

**Video S1. Speech decoding alternating with reading in Bravo-3**

An online demonstration of speech-decoding alternating with reading in Bravo-3. The participant attempts to speak the target word on the screen at her own pace. After choosing to stop reading an article on the computer screen, the participant makes a speech attempt. This speech attempt is detected from neural activity and the corresponding neural activity during the speech attempt is passed to a speech-verification model. If the speech-verification model assigns a high probability to the event being attempted speech and not reading or listening, the neural activity is passed to a classifier that outputs the most likely word in the vocabulary. The system retains high-specificity to attempted speech without false positive activations during the reading portions of the task.

**Video S2. Speech decoding during the listening condition in Bravo-1**

The same decoding framework as described in Video S1; however, now the participant, Bravo-1, listens to a podcast and decides when to attempt to speak. The system again retains high-specificity to attempted speech without false positive activations during the listening phase of the task.

**Video S3. System specificity for attempted speech during listening in Bravo-1**

In this video, Bravo-1 listens to a podcast with the speech-decoding system, described in Videos S1 and S2, active. The speech-decoding system is not activated over several minutes of listening, maintaining high-specificity for attempted speech.
